# Appraising the HIV Prevention Cascade methodology to improve HIV prevention targets: Lessons learned from a general population pilot study in east Zimbabwe

**DOI:** 10.1101/2024.09.07.24310075

**Authors:** Louisa R. Moorhouse, Jeffrey W Imai-Eaton, Rufuworkuda Maswera, Blessing Tsenesa, Phyllis Magoge-Mandizvidza, Brian Moyo, Owen Mugurungi, Constance Nyamukapa, Timothy B Hallett, Simon Gregson

## Abstract

**Introduction:** Multiple HIV Prevention Cascades (HPC) formulations have been proposed to assist advocacy, monitoring of progress of HIV prevention implementation and research to identify ways to increase use of HIV prevention methods. Schaefer and colleagues proposed a unifying formulation suitable for widespread use across different populations which could be used for routine monitoring or advocacy. Robust methods for defining and interpreting this HPC formulation using real world data is required.

**Methods:** Data collected as part of the Manicaland Pilot HIV Prevention Cascades Study, east Zimbabwe, in 2018-19, was used to validate the HPC framework for PrEP, VMMC, male condom and combination prevention method use. Validation measures included feasibility of populating the HPC, contrasting simple vs complex measures of the HPC (using 2-sample proportion test), and testing ability of main bars to predict prevention use and testing whether sub-bars explained why people were lost from the HPC using logistic regression.

**Results:** It was possible to populate the HPC for both individual and combined prevention methods using pilot survey data. Most steps were associated with prevention method usage outcomes, except for VMMC. There were significant overlaps between individuals reporting positive responses for the main bar and those citing barriers to motivation. To refine the HPC’s access bar definition, it is suggested to also consider individuals who report access barriers. While the HPC framework identifies barriers to individual prevention methods, challenges arise in identifying those for combined prevention.

**Discussion:** Our study successfully utilised questionnaires from the Manicaland HPC pilot survey to measure the HPC for individual and combined prevention methods. This demonstrates the feasibility of populating this framework using general population survey data and designated questionnaire modules. We propose a final formulation of the HPC, questionnaire modules and methods to create it. With proper evaluation and promotion, the HPC can enhance prevention services, aiding in the crucial reduction of HIV incidence.

## Introduction

Following the success of the HIV Treatment Cascade framework to compare and evaluate national and sub-national HIV treatment programmes1–3, multiple models of HIV prevention cascades (HPC) have been proposed to monitor progress of HIV prevention implementation from national to local levels, identify ways to increase use of HIV prevention methods, allow comparison across HIV prevention programmes, and ultimately reduce HIV incidence^4–11^.

### The intended use of the HIV Prevention Cascade Framework

The aim of the HPC model is to provide a practical framework that indicates where HIV prevention activities need to be strengthened by describing the steps required for HIV prevention to be effectively used by an individual, and identifies barriers to the individual transitioning through each of these steps^5,6,12^. Successful application of the HPC must be able to assist decision makers to identify intervention uptake targets that result in improved effective use of HIV prevention^6,13^.

Garnett et al proposed two general models of an HPC which were applied to data from the Manicaland cohort study in Zimbabwe^5^. One of these HPCs takes the view of a health care provider, i.e., provider-centric and one which takes the view of an individual potentially engaging with an HIV prevention method i.e., user-centric^5^. These were the first of the proposed HPC models which were more generic: rather than being developed for a specific population, such as MSM, or prevention method, such as PrEP. Hargreaves and colleagues modified and built on the HPC formulations proposed by Garnett et al^5^. First, by identifying three domains of the cascade that interventions could be related to: demand side interventions which aim to increase motivation; supply side interventions which improve availability; and adherence interventions to improve uptake and use of HIV prevention. Second, by incorporating known barriers in each of these domains drawn from the earlier social cognitive theoretical frameworks and the wider literature and linking these to the types of interventions most likely to reduce these barriers^12^. i.e., in effect, an explanatory framework.

However, such theoretical frameworks were not designed for use in routine programme monitoring and advocacy and there was a need for a unifying framework which could be used for such work.

Following a consultation in Harare on earlier HPC formulations, Schaefer *et al*. and the London Working Group on HIV Prevention Cascades (LWG) proposed a unifying cascade framework for routine monitoring and evaluation of prevention programmes^6^. It also highlights gaps between the key steps that need to be addressed to achieve effective prevention method use. The framework can be applied to multiple primary prevention methods and populations^6^. Schaefer *et al*. highlight that a successful cascade framework must be sufficiently generic to adapt across prevention methods and populations, and be efficient and practical to populate with real world data^6,14^. The resulting framework was proposed to be applied to multiple populations and to be used in two parts: a simple framework consisting of the main bars of the cascade (motivation, access, effective use) and a more complex framework which considers explanatory barriers to each of the gaps in the HPC (lack of motivation, lack of access, lack of capacity to effectively use)^14^. Recent studies have reinforced the importance of including individual-level motivation within the framework^14,15^. The inclusion of individual-level motivation distinguishes the Schaefer et al framework from other proposed frameworks such as that included in recent operational guidance from UNAIDS on creating HIV prevention cascades^16^. The UNAIDS approach adopts a more programmatic perspective with the cascade steps consisting of identifying a focus population and then measuring the reach/coverage, uptake/use and then correct/consistent use of primary HIV prevention methods^16^.

It is possible that, as suggested by Auerbach *et al*., two different cascade formulations are required— one simple model which populates the core steps, and thus highlights gaps in the cascade, to evaluate prevention programmes, complemented by a more detailed model which includes explanatory factors for the gaps observed in the cascades^17^. This is similar to the approach suggested by Schaefer *et al*. whereby the core steps of the cascade framework are populated initially and then, if data are available, explanatory sub-bars are populated as reasons underlying the gaps in the cascade^6^. However, the utility of any version of the cascade framework, either simple or extended, needs to be demonstrated using data to specifically populate HPCs for both individual and combination HIV prevention^18^.

### Collection and application of real-world data to populate the HIV Prevention Cascade

Preliminary research has demonstrated populating the HPC using pre-existing general population survey data^5,6,14,19^ ; however, relying on pre-existing data has been recognised repeatedly as a limitation, particularly for populating the explanatory barriers. Several studies applying the LWG/Schaefer version of the HPC to various priority populations have successfully populated the core steps of the HPC cascade^20^, but these studies have reported lack of data availability when using the extended framework with explanatory sub-bars^20–22^.

Despite an emerging consensus that the cascade should consist of initially defining a priority population, followed by measuring motivation, access and use within this population, data collected specifically to populate and validate the cascade framework are lacking^18^. Before encouraging use of this HPC framework, it is necessary to understand and demonstrate the practicality of collecting data to specifically populate the cascade. The Manicaland Study, an ongoing open cohort study in east Zimbabwe, has collected data to demonstrate the practical utility of the particular generic HPC proposed by LWG/Schaefer *et al*^6^. The pilot survey tested a questionnaire module to capture data on HPCs across multiple primary HIV prevention methods: male condoms, female condoms, PrEP and VMMC.

### Validating the HIV Prevention Cascade Framework

The LWG/Schaefer HPC framework requires validation using data specifically collected to populate the framework. Reviewing the validity of the proposed HPC framework aims to assess whether the requirements of its specific intended use are fulfilled when populated with survey data. As emphasised repeatedly, a key feature of any cascade formulation is that it must be simple and practical to populate^5,6,14^. Therefore, the most parsimonious version of the HPC framework possible should be favoured when reviewing the validity of measures and survey tools to populate the framework whilst trying to maximise the information which can be gained from the framework. This will maximise the likelihood of the HPC being adopted across national and sub-national programmes and research, as well as being the most feasible to include in population surveys such as Demographic Health Surveys. Collecting data via population surveys is crucial to understand the full picture of HIV prevention method use within the larger population compared to clinic-based programmes.

Defining the priority population, the denominator of the HPC framework, is the first challenge to populating the cascade. Specifying a priority population for prevention is more complex than the definition of a starting population of the HIV treatment cascade^17^. The definition of the initial denominator of the population at risk will have a knock-on effect on the entire cascade^17^. Schaefer *et al*. define the priority population as the “population that could benefit from using a prevention method”, a definition which is open to adaptation according to the (national or sub-national) population of interest^6^. Within the context of Manicaland, work has been done to establish the definition of priority populations through a combination of literature review and analysis of sexual behaviours associated with HIV acquisition^6,23^.

Following the definition of the priority population, the LWG/Schaefer HPC core framework consists of three main bars: motivation, access, and effective use of prevention methods^6^. Motivation captures an individual’s desire to use a prevention method. Access captures whether an individual is able to access a prevention method. Effective use describes the use of a prevention method required to avert acquisition of HIV^6^. For the proposed HPC framework to successfully describe the steps taken for an individual to effectively use HIV prevention, the steps leading to effective use must be predictive of effective use. Although the core cascade is designed to be generic and applicable across populations, validating the questionnaire module developed to populate the framework and the combinations of questions to measure each domain of the cascade is required to help promote use of the HPC framework.

Schaefer et al. stress the importance of individual-level motivation in prevention method use and therefore suggest that this should be the first step in the HPC^6^. Early exploratory analysis of the cascade has consistently indicated a very small drop between the motivation bar and the access bar. This raised questions about how the main bar for access was being defined and whether using a single question – “do you know a place you could access a prevention method” – was sufficient and identified a need to explore the overlap between motivation to use and access to prevention methods. This compounds a point raised during the Harare workshop that the HPC may not be as clearly linear as the treatment cascade, meaning the proposed order of motivation and access may be reversed or highly correlated^14^. This hypothesis requires testing using general population data. Schaefer *et al*. highlight that, given that both motivation and access are necessary for effective use, their order in in the cascade is unlikely to impact programmatic decisions. Regardless of access, an individual will not use a prevention method if they are not motivated to, and individuals may still experience barriers in their capacity to use prevention methods effectively^6^. However, if interventions to improve HIV prevention methods are designed based on gaps identified in the cascade then the HPC framework must effectively highlight these gaps – something which could be affected by the order of the bars^13^. Auerbach *et al*. also suggested that in priority populations which have very high coverage of access to a prevention method, ordering the cascade so that access comes first would give greater insight to motivation to use a prevention method when access is not an issue^17^. The different insights gained from the HPC framework when swapping the order of motivation and access however remain to be demonstrated.

The extended version of the LWG/Schaefer HPC framework includes sub-bars which act as explanatory variables hypothesised to explain the gaps in each bar of the cascade. The sub-bars apply to specific domains of the cascade and therefore it is hypothesised in the framework that these explanatory variables are associated with a lack of each domain. For example, explanatory variables hypothesised to explain a lack of motivation should be associated with a lack of motivation. Inclusion of these variables in the framework was based on extensive literature review and models of behaviour change^12^, but it is necessary to test these associations using data specifically collected to populate the HPC framework^6,24^.

Further to work that has been carried out so far, robust methods for defining and interpreting HPCs are required. It is necessary to validate:

1. The steps and sub-bars in the cascade framework
2. The questionnaire module developed to populate and measure the cascade framework

We aim to assess and refine the ability of the HIV Prevention Cascade and questionnaire module proposed by Schaefer et al. to identify potential targets for HIV prevention interventions using data from a general population survey through the following objectives:

1. Test the feasibility of populating the HIV Prevention Cascade draft formulation with questions in the prevention questionnaire tool collected in the Manicaland pilot survey
2. Test whether the motivation and access main bars of the cascade frameworks are predictors of effective use of HIV prevention methods
3. Contrast alternative ways of measuring each main bar in the cascade defined in the Manicaland HIV Prevention Cascade pilot survey questionnaire module
4. Compare the populations captured within each main bar of the HPC draft formulation when the order of the motivation and access bars are swapped
5. Test the validity of the sub-bars to explain why individuals are lost from the HPC and are not effectively using HIV prevention methods
6. Propose a final validated version of the HPC framework
7. Propose a minimum questionnaire module to populate the main bars and explanatory sub-bars of the HPC framework

## Methods & Materials

### Data sources and study setting

Data were from the Manicaland HIV Prevention Cascade Pilot Study collected in 2018-19, which was carried out in Manicaland Province in eastern Zimbabwe^25^. Data on sociodemographic characteristics, HIV knowledge, risk and prevention method use were collected in a pilot questionnaire designed specifically to populate HPCs^26^. Ethical approvals for all survey activities were granted by the Medical Research Council of Zimbabwe and the Imperial College Research Ethics Committee (17IC4160).

### Manicaland HIV Prevention Cascade Questionnaire module

The Manicaland HPC pilot study implemented a draft questionnaire module containing questions to populate HIV prevention cascades for male condoms, female condoms, VMMC, PrEP and HIV testing^26^. Questions were proposed following a stakeholder consultation during the Harare Workshop^14^ and developed into HIV prevention modules of an individual questionnaire^14^. As part of this pilot survey, PrEP adherence laboratory testing was conducted on a sub-sample of young females reporting current or recent PrEP use^23,27^. Current PrEP adherence was defined by a concentration of Tenofovir above 0.7pM per DBS punch^28^.

### Definition of the priority population

To maximise the sample size available for validation purposes, a broad definition of a priority population was used: HIV-negative participants aged 15-54 years who reported one or more sexual risk behaviours for HIV acquisition in the last 12 months. HIV status was determined by either PITC or laboratory-based testing of DBS specimens^23^. Sexual risk behaviours were selected based on literature review and an HIV incidence analysis of the Manicaland general population cohort^23^. Risk behaviours included were having multiple partners in the last 12 months; concurrent partners at the time of interview; recent transactional sex in the last month with any of the last three partners; and having at least one non-regular partner in the last 12 months.

### Populating the HIV Prevention Cascade

Definitions of each bar and sub-bar of the HIV prevention cascade for each prevention method are listed in Tables S1-S3.

Outcomes were coded as binary variables and the bar height was calculated as this proportion within the target population:

- 0 = does not meet criteria for that bar/sub-bar
- 1 = does meet criteria for that bar/sub-bar

For any main bar (being motivated to use, having access to or effectively using a prevention method) this was coded as:

- 0 = individual is not motivated to use a prevention method
- 1 = individual is motivated to use a prevention method

For barrier bars (such as knowledge/lack of knowledge as a barrier to prevention method use) this was coded as:

- 0 = individual does not experience a lack of knowledge as a barrier to prevention method use
- 1 = individual does experience a lack of knowledge as a barrier to prevention method use

An individual was only exposed to a barrier bar if they did not meet the criteria to fall within the main bar, so the cascade takes the perspective of those who have the positive motivation/access/effective use and then seek to understand the barriers for those who do not. It was assumed that all those who effectively use a prevention method were also motivated to use and had access to each HIV prevention method. The HPC framework was populated as a conditional cascade; each step was conditional on the previous step and therefore an individual must be in the previous step of the cascade in order to be allowed to experience the following step. For example, an individual must be motivated to be considered in the denominator for the access bar.

Each main bar of the cascade was calculated as a proportion:

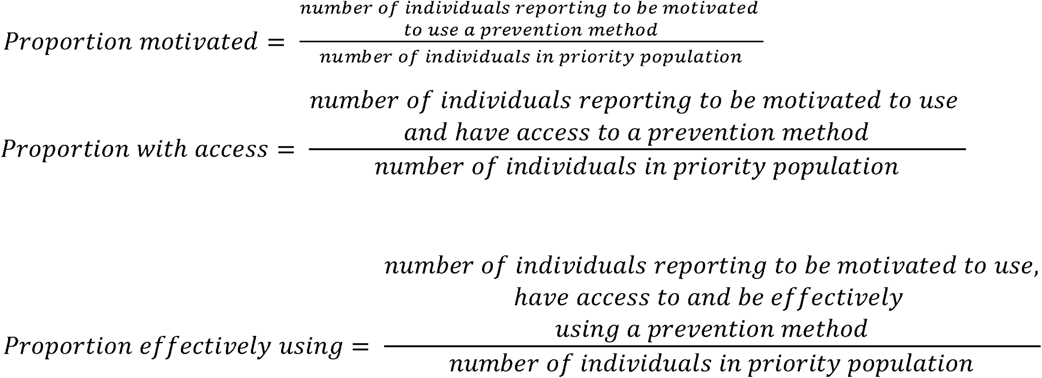

95% confidence intervals for each main bar proportion were calculated for the main bars of the HPC framework.

Explanatory sub-bars were populated for individuals who met the conditions to continue through to that domain of the cascade, but did not report the main bar for this domain positively. For example, to be in the access bar individuals must report to be motivated. Explanatory barriers for motivation were populated for those in the priority population but who were not motivated; for access, those who were motivated but lacked access; and, for effective use, those who were motivated and had access but did not effectively use a prevention method.

### Combination prevention method use

The combination HPC framework was populated by classifying whether individuals in the priority population used at least one of the prevention methods:

a. Males – VMMC and/or male condoms
b. Females – PrEP and/or male condoms

When calculating the main bars (e.g., motivation), individuals had to report affirmatively for those bars for at least one prevention method. This calculation built on definitions of individual prevention method use and so the combination cascade remained conditional specific to individual prevention method use e.g., an individual could not be part of the combination motivated bar based on male condom motivation and then part of the combination access bar based on VMMC access.

Individuals were included within combination explanatory sub-bars when they did not fall into a main bar (e.g., motivation) for any prevention method. The sub-bars were assigned to the furthest possible point along the cascade, with the end point of any prevention method use in mind. For example, if an individual was motivated to use male condoms but lacks motivation to use VMMC, they continued to the access domain of the framework because they were motivated to use at least one method. At this point, if they reported a lack of access to male condoms, then explanatory barriers to this access should be reported within the cascade.

### Measuring validity of the HPC framework

Hypotheses tested within each objective are listed in Table 1. Accepting or rejecting each hypothesis is not solely based on statistical outcomes but also on the practicality and feasibility of each part of the HPC framework, and considering the previous justifications for each part of the HPC framework as set out by Schaefer et al, the LWG and during the Harare Workshop.

**Table 1 -.**
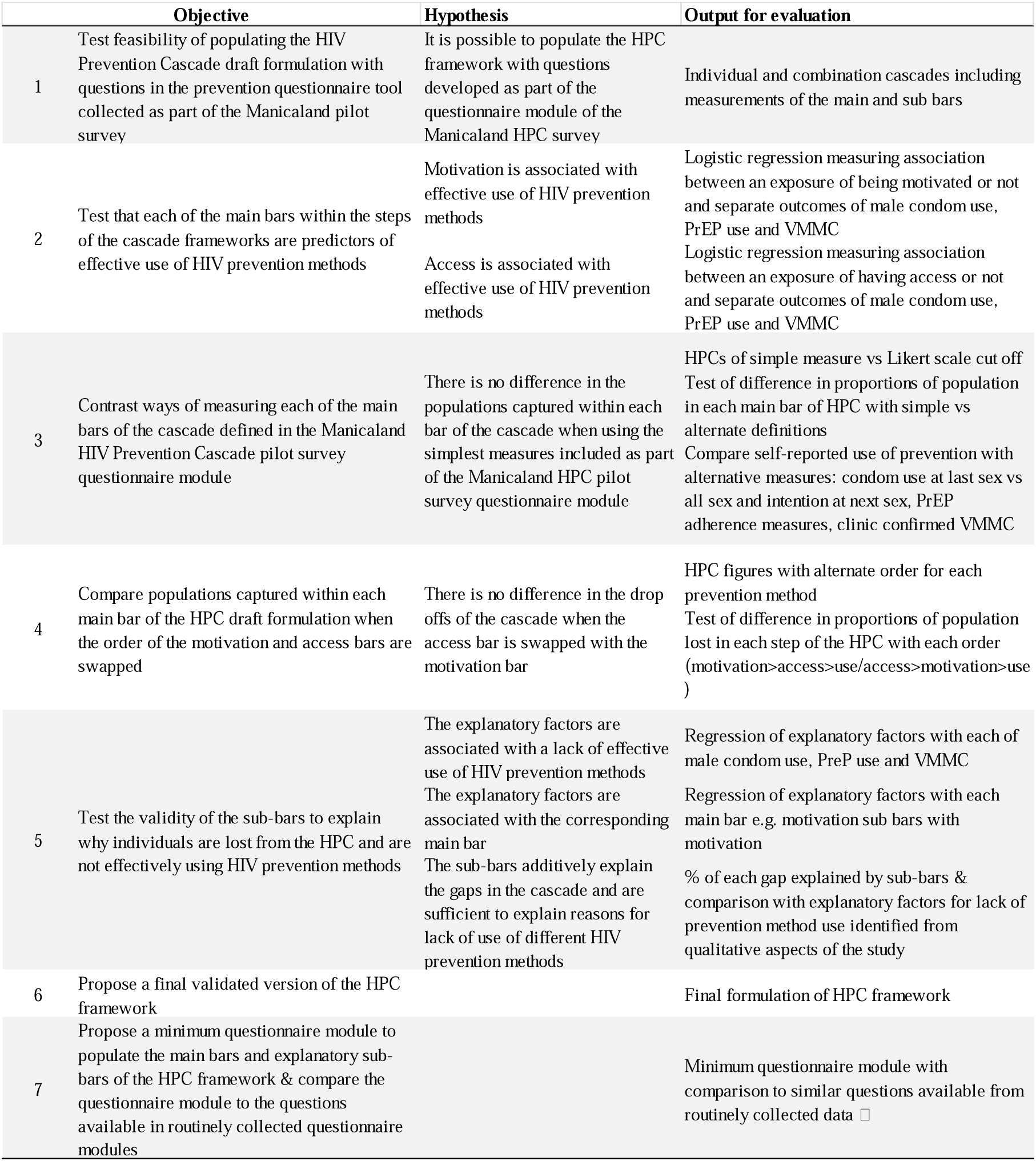
Hypotheses tested within each objective of the validation process and the output evaluated within each objective.

#### Objective 1

The feasibility of populating the HPC framework was tested using data collected from the Manicaland pilot questionnaire modules. Full cascades were populated for male condoms, PrEP and combination prevention in the female priority population and male condoms, VMMC and combination prevention in the male priority population.

#### Objective 2

Logistic regression was used to test the association of motivation and access as predictors of the outcome of effective use of HIV prevention methods. Regressions were fitted separately for each prevention method (male condoms, PrEP and VMMC), adjusted for 5-year age group and site type, and stratified by sex. Statistical significance was assessed using a threshold of p<0.05.

#### Objective 3

Multiple potential questions to populate each main bar of the HPC framework were included in the Manicaland pilot questionnaire module. The simplest combination of questions used to populate the cascade was considered the most favourable measure to ensure the HPC framework is as feasible as possible to routinely collect data on. Each of the main cascade bars were calculated using the simplest (primary) measure and then also using more complex (alternate) measure, as detailed in Tables S1-S3, for each prevention method and stratified by sex. Throughout this analysis, the definition of the main cascade bars included those effectively using each prevention method. Differences in the proportions captured within each bar of the cascade using simple versus alternate measures were compared using a two-sample proportion test and p-values for two-tailed tests of significance between the proportions were used to assess the hypothesis that there is a difference in proportions identified through the simple versus alternative measure. Statistical significance was assessed using a threshold of p<0.05. Self-reported use of prevention methods was compared with alternative measures—self reported condom use at last sex versus condom use throughout all sex in the last year, self-reported PrEP use versus laboratory confirmed PrEP use and self-reported VMMC versus clinic confirmed VMMC^23^.

Logistic regression was used to test the association between these measures of prevention method use.

The proportion of individuals with overlap between reporting a positive bar and also reporting barriers to each domain of the cascade were calculated and presented using Venn diagrams e.g., those who report to be motivated to use a prevention method but also report barriers to motivation.

#### Objective 4

Cascades were populated for each prevention method, stratified by sex, using the alternate order of the main bars (access > motivation > effective use). The overall proportions lost from the cascade between the starting point of the priority population to the effective use bar with each order of the bars were compared. The proportions lost at each step of the cascade were compared for each order of the main bars using z-tests.

#### Objective 5

The association of explanatory factors with use of each prevention method was tested using multivariate logistic regression, adjusted for 5-year age group and site type, and stratified by sex. The association of each explanatory factor with each domain of the cascade was tested using multivariate logistic regression, adjusted for 5-year age group and site type, separately for each prevention method and stratified by sex, to test if the explanatory factors correctly explained the gap in the cascade they have been assigned to in the proposed HPC framework. The total proportion of each gap explained by the explanatory sub-bars was measured. Where the gaps were not fully explained additively by the explanatory sub-bars, the results were compared with explanatory factors for lack of prevention method use identified from individual interviews and focus groups from qualitative parts of the study and factors identified from qualitative work but not covered by the HPC framework were reviewed^23^.

#### Objectives 6 & 7

Based on results from these analyses, a final version of the HPC framework was proposed. A questionnaire module was recommended based on the minimum questions required to populate a simple (main bars only) and extended (including explanatory sub-bars) HPC framework. The simplest measures were favoured where possible to maximise the feasibility of use of the HPC framework across multiple settings and data sources including population level surveys.

All statistical analyses were carried out using Stata/MP 17.0. Data visualisation was carried out using Tableau.

## Results

9803 individuals aged 15 years and above completed the individual questionnaire (females n = 5729; males n = 4074), 77% of those eligible for participation from the household census. 9339 (95%) of the individuals completing the individual questionnaire had an HIV result, either from PITC (n = 7715) or DBS laboratory testing (n = 1624). 8404 individuals (90%) of those with an HIV test result were HIV negative. 6307/8404 (75%) self-reported sexual debut. 5223/6307 (83%) were aged 15-54 years.

Among those sexually active 15–54-year-olds, 14% of females and 28% males reported ≥1 HIV sexual risk behaviour giving priority populations of 575 and 444 HIV negative males and females respectively.

### Objective 1 - Test feasibility of populating the HIV Prevention Cascade draft formulation with questions in the prevention questionnaire tool collected in the Manicaland pilot survey

It was possible to create HPCs for PrEP and male condom use among females (Figure 1) and VMMC and male condom use among males (Figure 2) in the priority population using data collected within the Manicaland pilot questionnaire module. Levels of motivation, access and use of each prevention method could be assessed for each priority population and compared across prevention methods. Of women in the priority population, 63% were motivated to use, 62% were motivated and had access to and 32% reported motivation, access, and male condom use. PrEP use was lower: 10% were motivated to use, 8% had access to and <1% reported PrEP use. 52% of men in the priority population were motivated to have VMMC, 46% reported access and 19% reported full medical male circumcision. Male condom use rates by men was higher than VMMC: 74% were motivated to use, 74% had access to and 60% reported male condom use.

**Figure 1 -.**
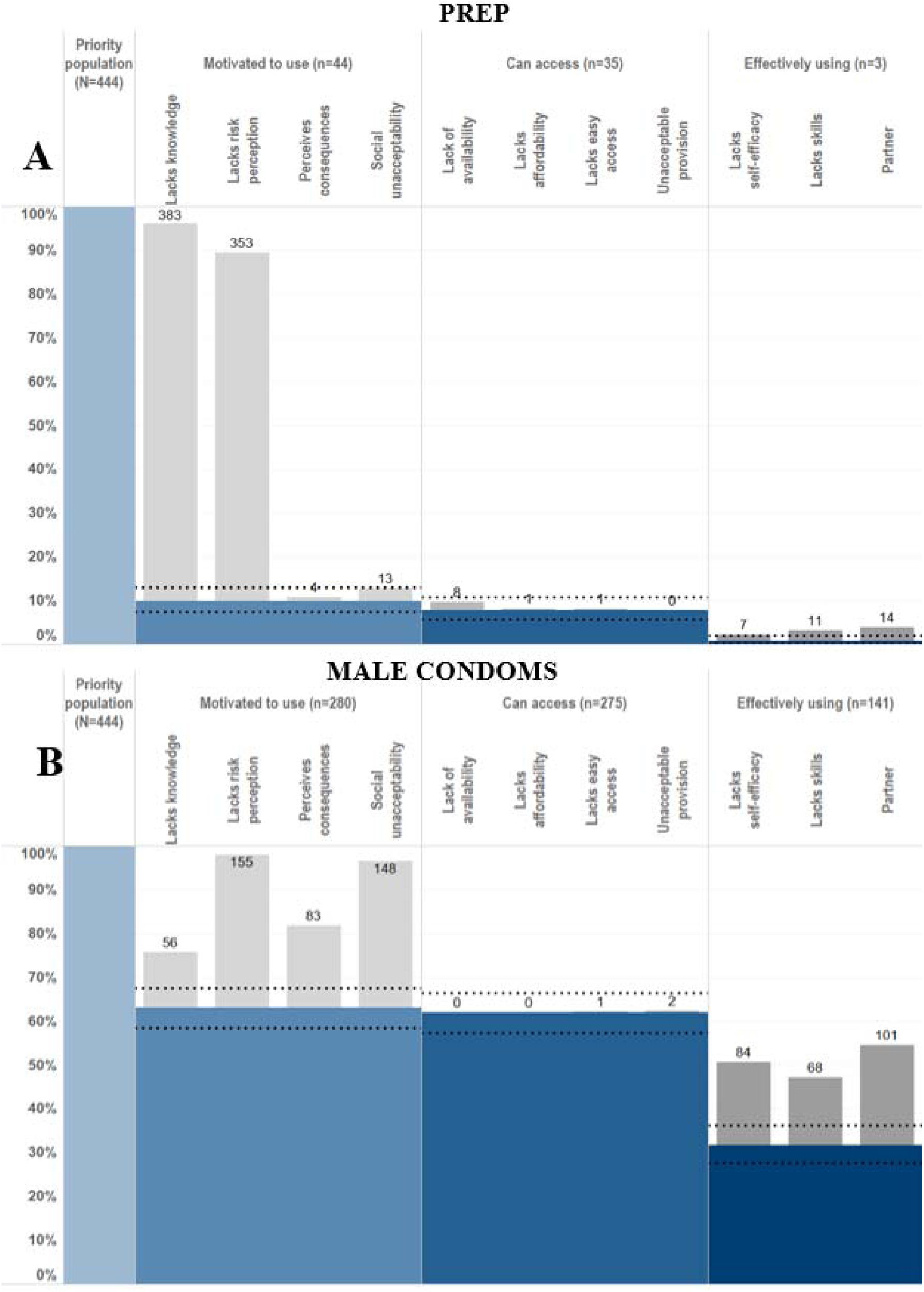
HIV prevention cascades for PrEP and male condom use in females in the priority population

**Figure 2 -.**
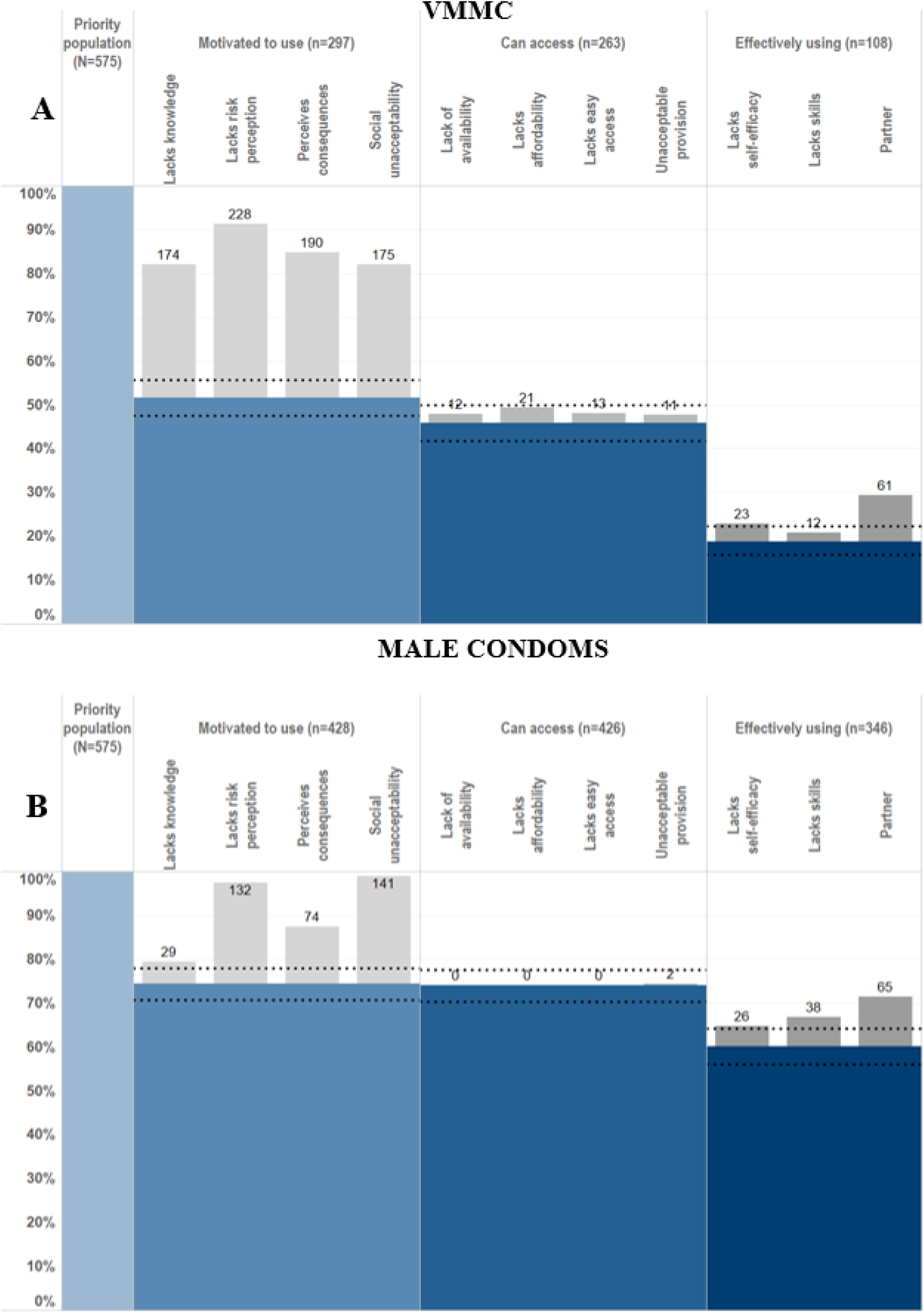
HIV prevention cascades for PrEP and male condom use in females in the priority population

Where gaps in the cascade were identified, it was possible to populate sub-bars explaining the gaps in the cascade and barriers to progressing through the cascade. Barriers observed include lack of knowledge, lack of risk perception, perceived consequences of use and partner resistance. It was also possible to populate combination prevention cascades (Figure 3). 67% of men and 32% of women reported using at least one prevention method. It was possible to populate the sub-bars to understand explanatory barriers to use although it was not possible to see which prevention method the barriers were relating to, which prevention methods were preferred, or the proportion of people using multiple prevention methods. Some barriers, such as a lack of risk perception and a lack of knowledge of all prevention methods, could still provide some insight into barriers to combination prevention method use.

**Figure 3 -.**
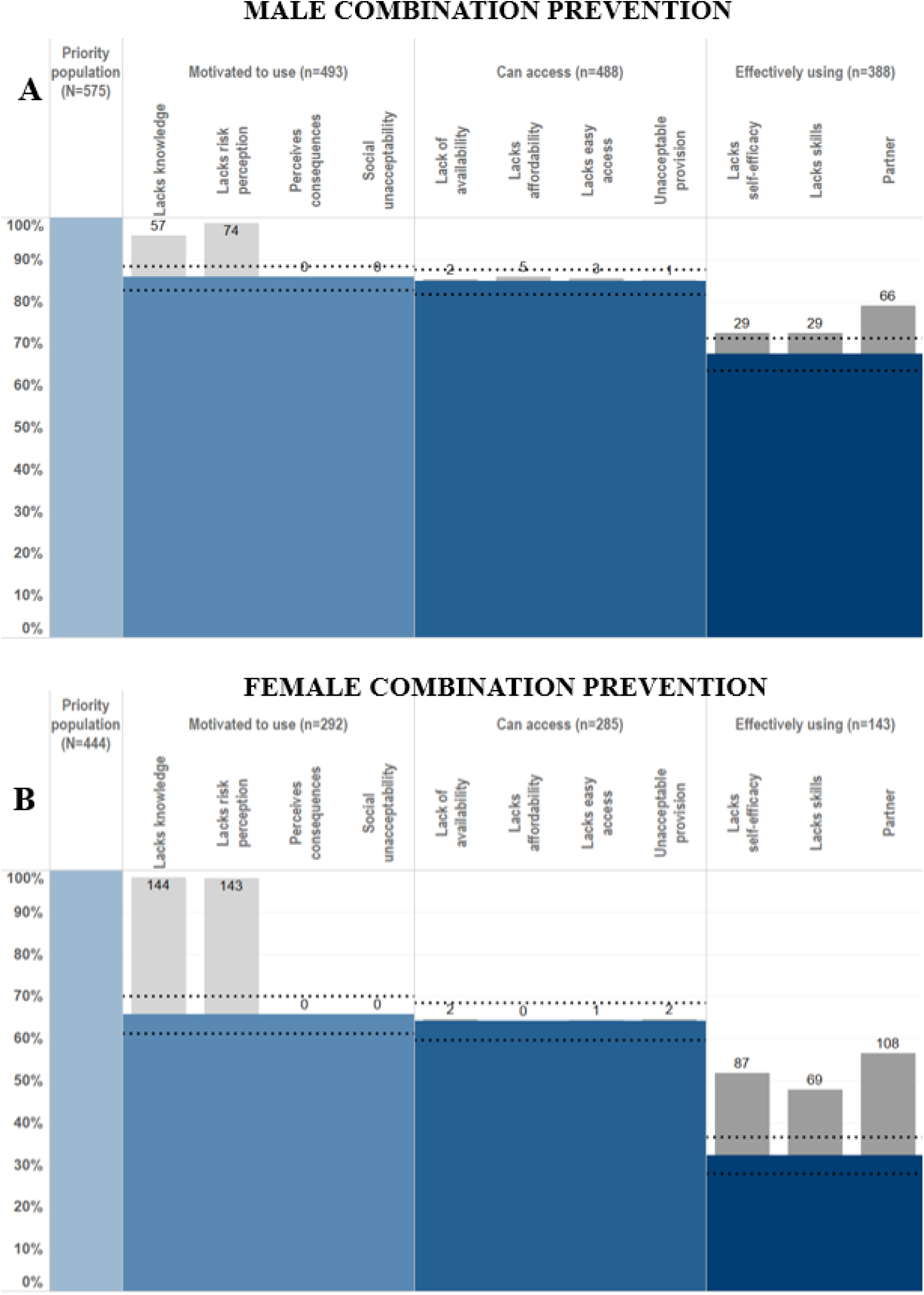
– HIV prevention cascades for HIV combination prevention: use of VMMC or male condoms in males and PrEP or male condoms in females

### Objective 2 - Test that each of the main bars of the cascade framework are predictors of effective use of HIV prevention methods

Logistic regression models, adjusted for site type and 5-year age-group, were used to measure the association between motivation and prevention method use, then access and prevention method use (Table 2). Among both women and men, motivation to use condoms and access to male condoms were strongly associated with use of male condoms, indicating that these are necessary components of the steps of the HIV prevention cascade. For PrEP, it was not possible to calculate odds ratios for associations of motivation and access with PrEP use due to small numbers of people in the priority population reporting PrEP use, all of whom reported motivation and access. Motivation for and access to VMMC were significantly associated with lower odds of having VMMC. This is likely due to the way the questions were asked: 95% of people reporting VMMC responded that they were not motivated to get VMMC or did not know somewhere to access VMMC if they did want to access it, probably because they had already had the procedure. If the HIV prevention cascade was populated without the ‘effective use assumption’—i.e., all reporting prevention method use must be motivated and have access to that prevention method—then inaccurate levels of motivation and access would be captured in the cascade.

**Table 2 -.** Associations between motivation and access with prevention method use, adjusted for site type and 5-year age group.

|  | Motivation |  | Access |  |
| --- | --- | --- | --- | --- |
|  | n (%) | OR (95% CI) | n (%) | OR (95% CI) |
| PrEP (Female) | 44 (9.9) | *collinear* | 4 (0.9) | *collinear* |
| VMMC (Male) | 252 (43.8) | 0.06 (0.02-0.15) | 228 (39.7) | 0.09 (0.03-0.22) |
| Male condoms (Female) | 280 (63.0) | 26.83 (11.73-61.35) | 183 (41.2) | 17.90 (8.91-35.97) |
| Male condoms (Male) | 428 (74.4) | 8.36 (5.58-12.53) | 391 (68.0) | 8.33 (5.57-12.44) |

### Objective 3 - Contrast alternative ways of measuring each of the main bars of the cascade defined in the Manicaland HIV Prevention Cascade pilot survey questionnaire module

Simple versus alternative, more complex, ways of measuring the main steps of the HPC (Tables S1- S3) were calculated and the proportions of the population captured were compared using two-tailed two sample test of proportions (Table 3). There were no significant differences between the populations captured in the simple versus alternative measures for motivation or access across all prevention methods explored. It was not possible to compare VMMC use to an alternate measure due a lack of data on other ways of confirming VMMC. Two alternate ways of defining male condom use were tested: firstly, self-reporting using condoms at every sexual encounter for as long as an individual has been sexually active, and, secondly, self-reporting using male condoms throughout last sex. Significantly fewer (p<0.001) both men and women reported using the simple measure compared to using condoms at every sexual encounter in both men and women, with a decrease from 32% to 6% in females and 60% to 16% in males. Comparing the simple measure to the second option—using condoms at last sex—significantly fewer men reported effective use (p<0.001) with a decrease from 60% to 43%. There was no significant difference in the female populations captured (p=0.432). Self-reporting PrEP use was compared to a measure of using PrEP all or most days in the last month and no difference was observed in the populations of women captured (p=0.999). Testing for the presence of tenofovir and emtricitabine was carried out on DBS collected in young women reporting current or ever PrEP use across the study (Figure 4). All DBS samples had concentrations of Tenofovir above the threshold for PrEP adherence (0.7pM per DBS).

**Figure 4 -.**
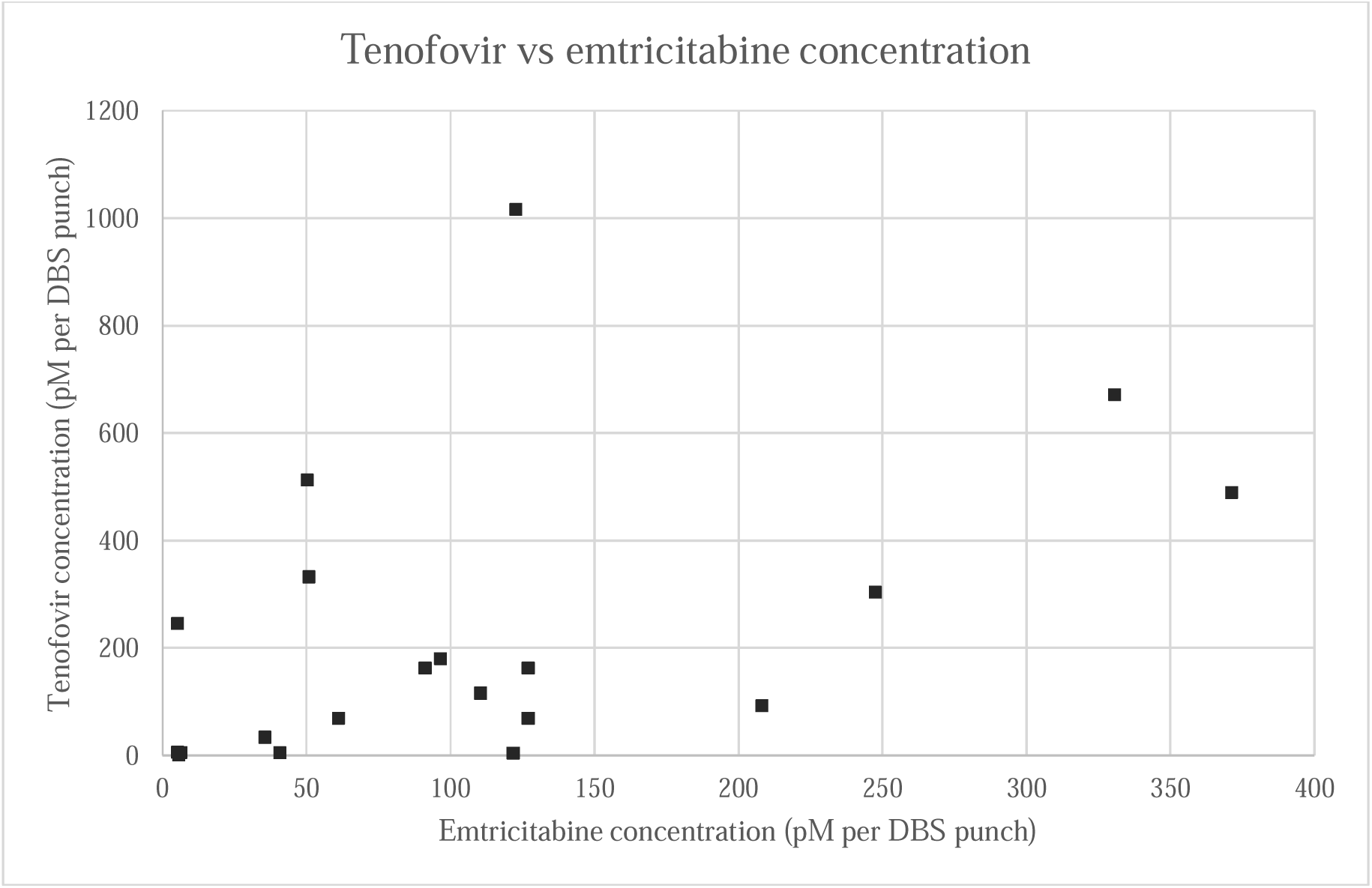
Tenofovir and emtricitabine concentrations from DBS testing in females aged 15-24 years reporting current or recent PrEP use

**Table 3 -.** Comparison proportions of motivation, access, and use of prevention methods with simple vs alternative measures and p-values for differences in proportions using 2-sample test of proportions.

|  | **Motivation** |  |  |
| --- | --- | --- | --- |
|  | **Simple measure** | **Alternative measure** | **p-value** |
|  | **% (95% CI)** | **% (95% CI)** |  |
| Male condoms - female | 61.5 (56.9-65.9) | 58.8 (54.1-63.3) | 0.411 |
| Male condoms - male | 63.5 (59.5-67.3) | 62.3 (58.2-66.1) | 0.669 |
| VMMC - male | 51.7 (47.6 - 55.7) | 51.0 (46.9 - 55.0) | 0.813 |
| PrEP - female | 9.9 (6.9-12.3) | 9.9 (7.5 - 13.1) | 0.999 |
|  | **Access** |  |  |
| Male condoms - female | 59.5 (54.8-63.9) | 57.2 (52.5-61.7) | 0.634 |
| Male condoms - male | 62.8 (58.7-66.6) | 62.3 (58.2-66.1) | 0.999 |
| VMMC - male | 27.8 (24.3-31.6) | 24.3 (21.0-28.0) | 0.179 |
| PrEP - female | 7.6 (5.1-10.0) | 6.8 (4.8-9.5) | 0.999 |
|  | **Use** |  |  |
| Male condoms - female | 31.8 (27.6-36.2) | 5.6 (3.8-8.2) | <0.001 |
| Male condoms at last sex* - female | 31.8 (27.6-36.2) | 34.2 (30.0-38.8) | 0.432 |
| Male condoms - male | 60.2 (56.1-64.1) | 15.8 (13.1-19.0) | <0.001 |
| Male condoms at last sex* - male | 60.2 (56.1-64.1) | 43.0 (39.0-47.0) | <0.001 |
| VMMC - male | 18.8 (15.8-22.2) | n/a | n/a |
| PrEP - female | 0.7 (0.2-2.1) | 0.7 (0.2-2.1) | 0.999 |

*\*Reported to have used male condoms throughout last sexual intercourse as measure of use of condoms*

Overlaps between populations reporting main bars (e.g., motivation) and reporting barriers to those populations were assessed for the whole priority population without the conditional nature of the cascade (Figures 5 & 6). 73% of men and 62% of women reported motivation to use male condoms but experiencing a barrier to use (Figure 5). However, 98% of people reporting use of male condoms also report a barrier to motivation. 23% of the male priority population and 11% of the female priority population report both access to male condoms and also barriers to access. 55% of males and 25% of females in the priority population report both use of male condoms and at least one barrier to male condom use. Conversely, for PrEP, women reported only small overlaps between motivation, access (Figure 6). Men reported larger overlaps in motivation to have VMMC and reporting a motivation related barrier (43%) (Figure 6). 11% reported both access and an access related barrier and only 4% reported to have VMMC and also experience a barrier to VMMC.

**Figure 5 -.**
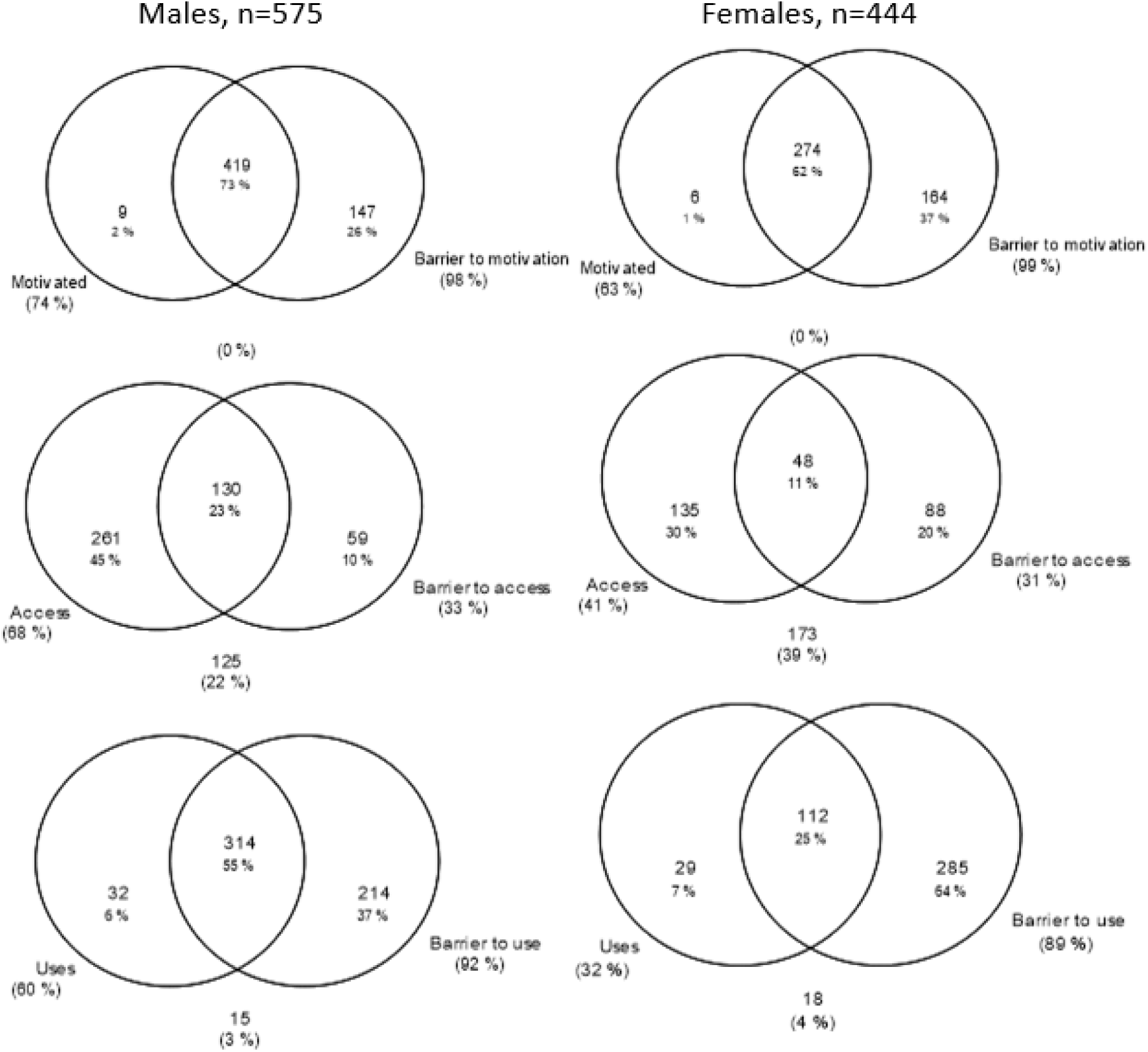
Venn diagrams of overlaps between each bar of the cascade and reporting at least one barrier to that bar for male condom use in males and females

**Figure 6 -.**
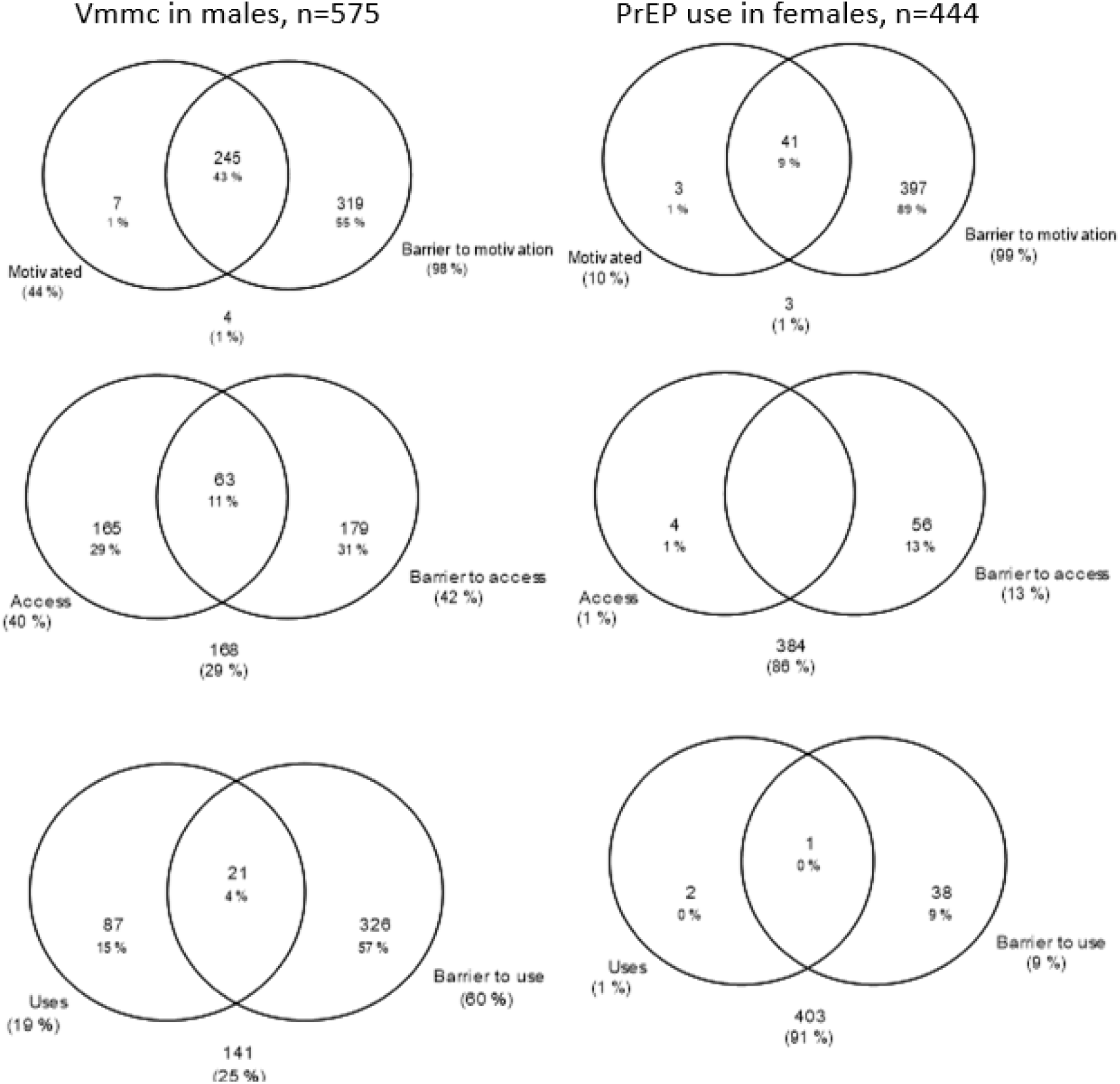
Venn diagrams of overlaps between each bar of the cascade and reporting at least one barrier to that bar for VMMC in males and PrEP use in females

### Objective 4 - Compare populations captured within each main bar of the HPC draft formulation when the order of the motivation and access bars are swapped

HIV prevention cascades were populated, for the main bars only, with the order of the main bars swapped and access being the first step of the cascade (Figures 7 & 8). The proportions of the population reaching the ‘use’ bar of the cascade were unchanged across all prevention methods. As with the first version of the cascade (Figure 1B), only a very small proportion of the priority population reported a lack of access to male condoms (5%) (Figure 7B). There was still a large gap in motivation with a large drop between the access and motivation bars: 34% of those with access were not motivated to use male condoms. Access to PrEP was low, and lack of access was the largest gap in the PrEP cascade in women, with 89% of the priority population lost from the cascade here. The drop was smaller between the access and motivation bars of the cascade with 66% of those with access reporting a lack of motivation to use PrEP (7% of the entire priority population).

**Figure 7 -.**
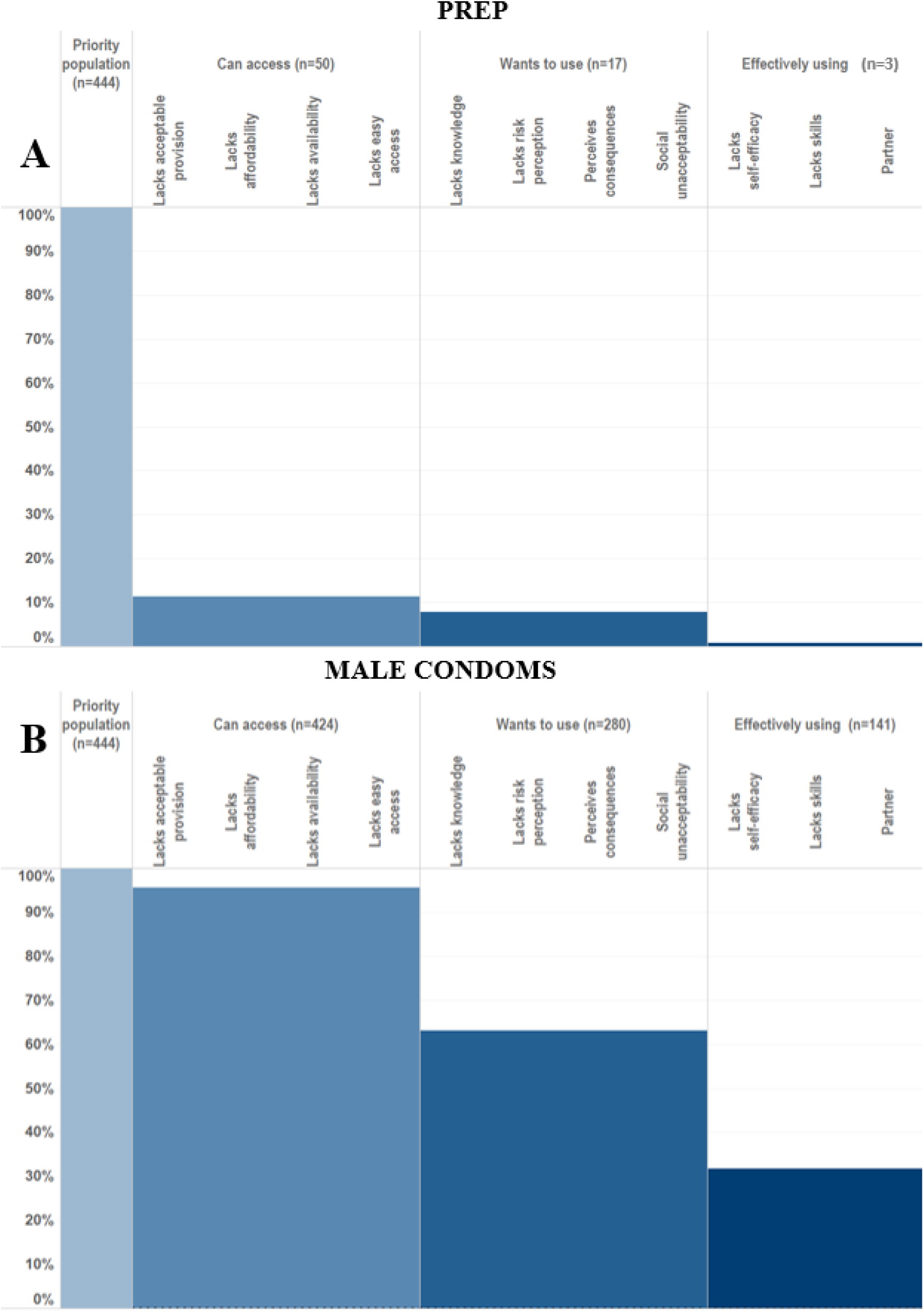
HIV prevention cascade with the order of Access, Motivation and Use, for PrEP and male condom use in females

**Figure 8 -.**
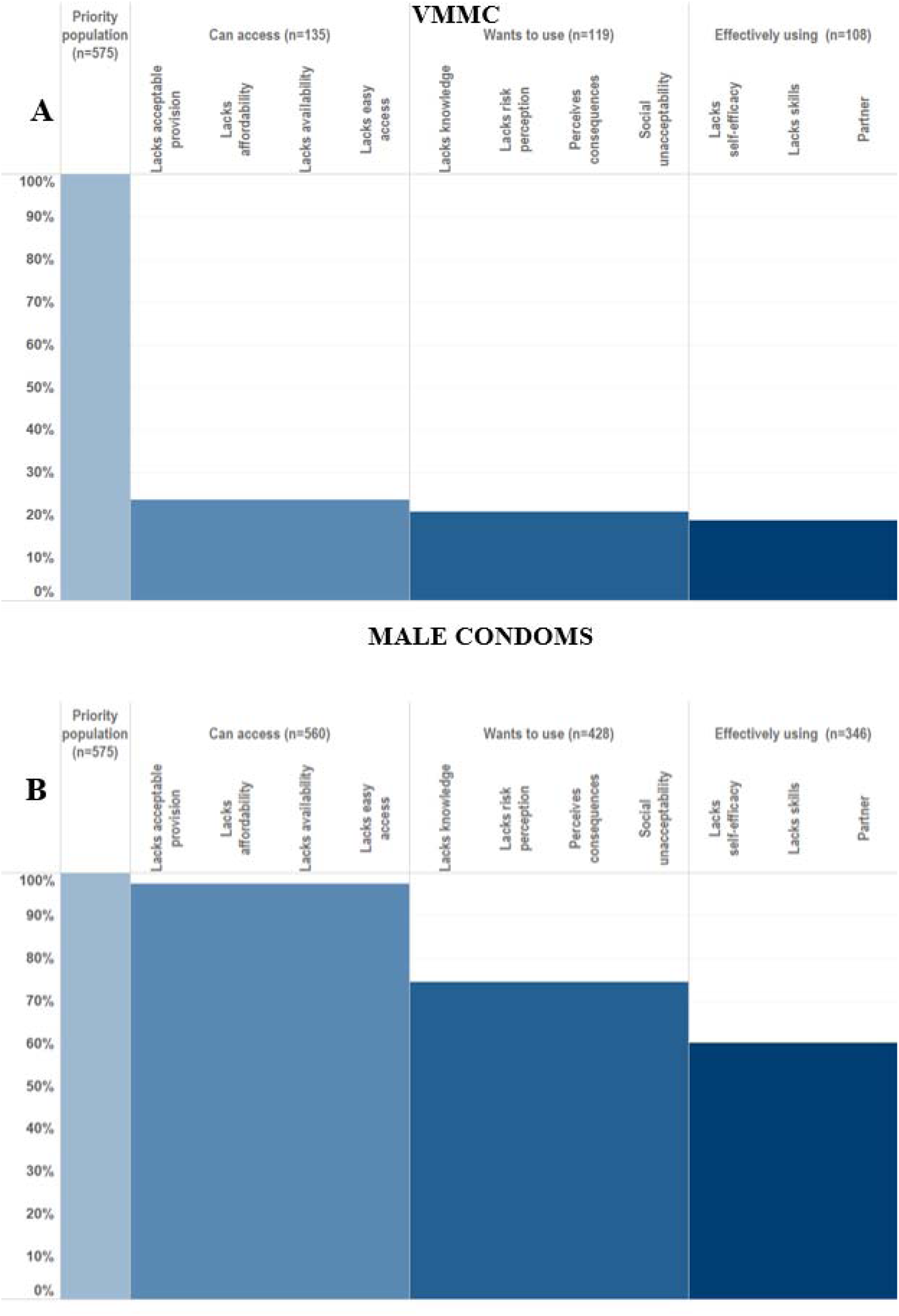
HIV prevention cascade with the order of Access, Motivation and Use, for VMMC and male condom use in males

77% of the male priority population reported a lack of access to VMMC (Figure 8A). The drop was small, 3%, between access to motivation. The drop was also small between the motivation and use bars: 2% of the total priority population. This is considerably smaller than the equivalent drops using the alternative order (Figure 3A) where 27% of the total priority population were lost from the second to third step. 2.6% of the male priority population reported a lack of access to male condoms (Figure 8B): a very small gap in access which is similarly observed using the alternative order (Figure 2B).

24% of men in the priority population with access to male condoms were not motivated to use them, representing 23% of the total priority population. 19% of men with access and motivation were not using male condoms. 14% of the overall priority population were lost from the motivation to use step here; the same proportion as lost from the access to use step in the original order of the HIV Prevention Cascade.

### Objective 5 - Test the validity of the sub-bars to explain why individuals are lost from the HPC and are not effectively using HIV prevention methods

Logistic regression models, adjusted for 5-year age group and site type, were used to assess the association between each of the hypothesised barriers in the HPC framework and use of each prevention method (Figure 9). Associations of PrEP related barriers with PrEP use were not calculated due to the very small numbers (n=3) reporting PrEP use.

**Figure 9 -.**
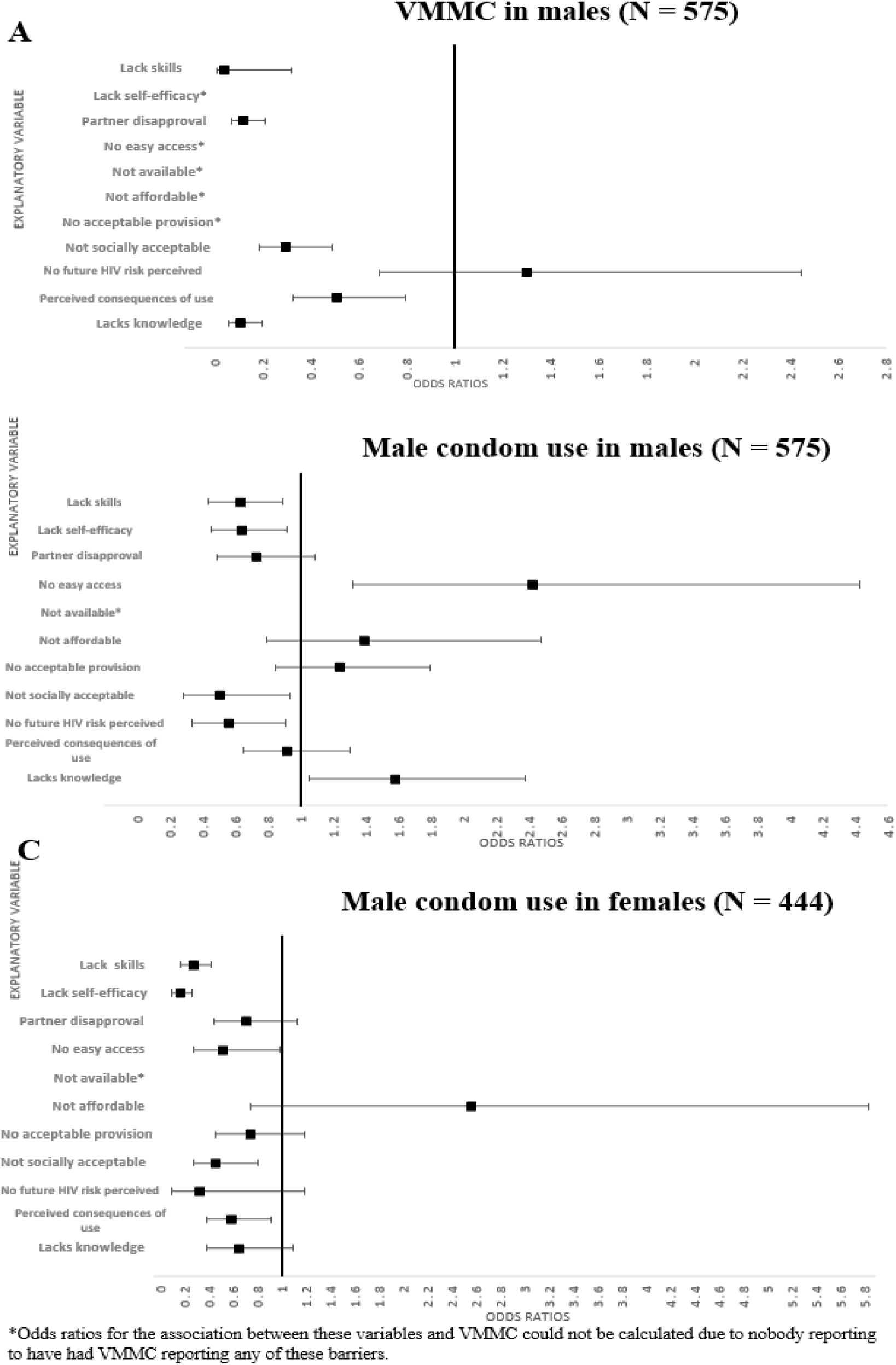
Associations of explanatory barriers with male condom use in females and male condom use and VMMC in males, adjusted for 5-year age group and site type

Odds ratios for the association of VMMC with lack of self-efficacy, easy access, availability, affordability, and acceptable provision could not be calculated because nobody reporting VMMC reported any of these barriers. Almost all other hypothesised barriers were significantly associated with lower odds of having VMMC (Figure 10A). Lack of skills and partner disapproval were significantly associated with reduced odds of having VMMC: OR = 0.04 (95%CI: 0.01-0.32) and OR = 0.12 (95%CI: 0.07-0.21), respectively. Lack of social acceptability (OR = 0.3 (95% CI:0.19-0.49), lack of knowledge (OR = 0.11, 95% CI: 0.06-0.20) and perceived negative consequences (OR = 0.51, 95% CI: 0.33-0.80) were all associated with reduced odds of VMMC among men in the priority population.

**Figure 10 -.**
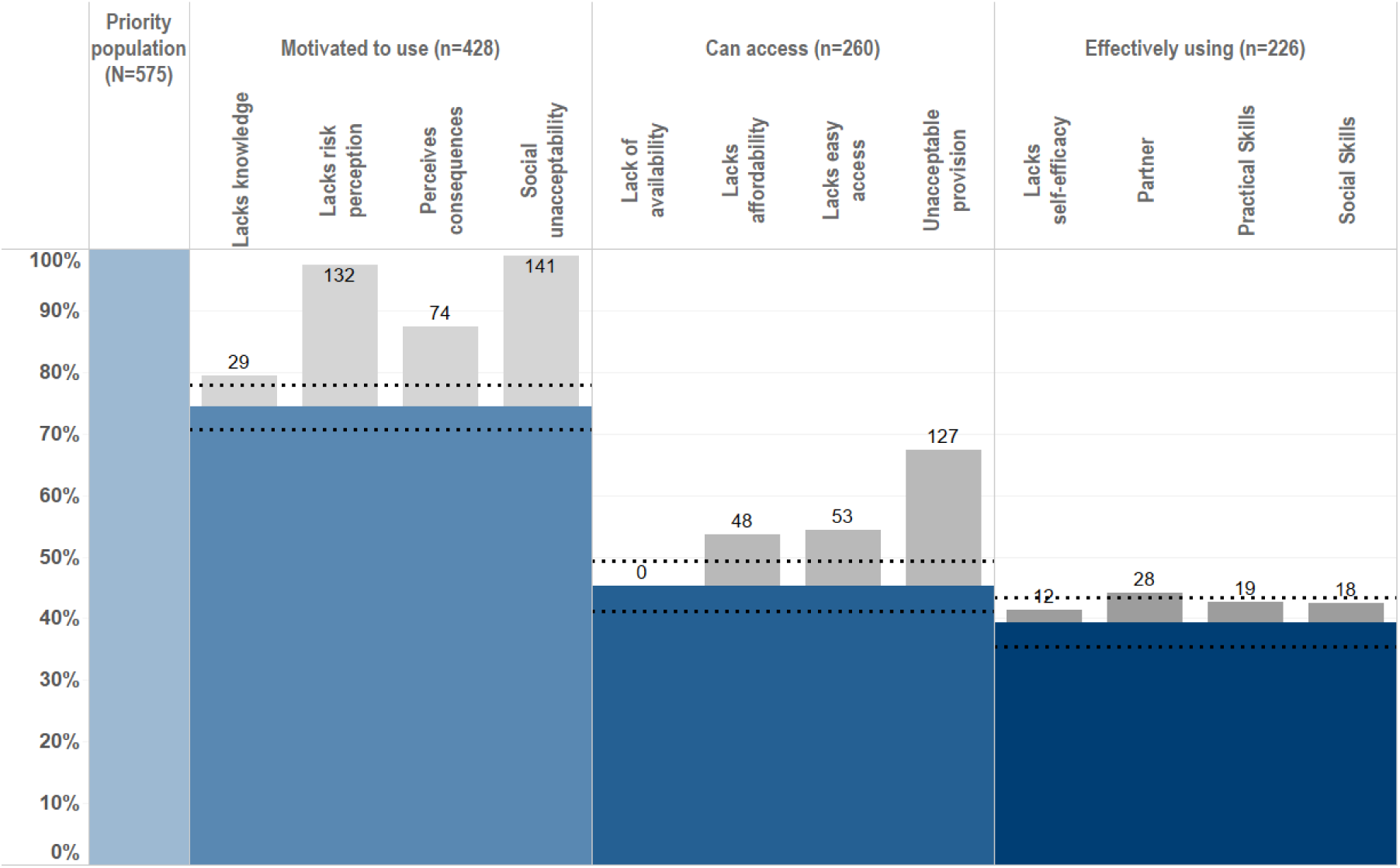
Final proposed formulation of the HIV Prevention Cascade, populated with data on male condom use in men

The only access related barriers significantly associated with lower odds of male condom use among men (Figure 9B) or women (Figure 9C) was lack of easy access in females (OR = 0.52 95% CI: 0.28- 0.98). Lack of skills and self-efficacy were associated with lower odds of male condom use in men and women. The motivation related barriers associated with lack of male condom use were different between men and women. In men, a lack of social acceptability (OR = 0.52 95% CI: 0.28-0.94) and a lack of future risk perception (OR = 0.56, 95% CI: 0.34-0.91) were the motivation related barriers associated with lack of male condom use. In women, a lack of social acceptability (OR = 0.46, 95% CI: 0.27-0.81) and perceived negative consequences of use (OR = 0.59, 95% CI: 0.39-0.91) were the motivation related barriers associated with lack of male condom use.

## Discussion

### Summary

This validation exercise demonstrated that it is feasible to populate the HIV prevention cascade framework, including both the main and sub bars, using data collected as part of the Manicaland Pilot HIV Prevention Cascades Study. The main bars in the HIV prevention cascade (motivation and access) predicted higher odds of prevention method use in male condoms and most sub-bars were associated with lower odds of prevention method use. High levels of overlap were observed of respondents reporting both the main bars and barriers to each main bar. Very little additional information was gained by swapping the order of the motivation and access bars of the cascade and in some instances, such as VMMC, this swap would lead to smaller gaps being identified and thus less insight being gained from the HPC framework.

### Strengths of the cascade framework

Using the questionnaire modules piloted in this study, it was possible to define the priority population based on questions on sexual risk behaviours for HIV acquisition and then assess the HIV prevention cascades for this priority population in men and women for both individual and combination prevention method use. Main bars of the cascades for both individual and combination prevention could be populated, giving insight into levels and gaps of motivation, access, and use of male condoms, VMMC and PrEP, as well as overall levels of VMMC and male condom cascades for men and PrEP and male condom cascades for women.

This demonstration of the feasibility of applying an HIV prevention cascade framework to combination primary HIV prevention is one of the first instances of such analysis^22^. In this analysis, the outcome of combination prevention considered was use of at least one prevention method.

Depending on the priority population and the research question, combination cascades could be constructed with an outcome of motivation to use, access to and use of 2 or more prevention methods together where multiple prevention methods are recommended in tandem to provide better protection e.g., VMMC and male condoms. Additionally, overall combination prevention cascades could be constructed in which all different combinations of prevention methods are considered. This version of the cascade framework allows this flexibility in application, provided the data are available.

Application of the framework to combination prevention was one of the reasons highlighted by Schaefer *et al*. in the need for a standardised framework^6^.

Motivation to use and access to male condoms were strongly associated with increased odds in condom use in both men and women, supporting the theory behind the inclusion of these key steps in the HPC framework. Due to the small numbers reporting PrEP use, motivation to use and access to PrEP were collinear with PrEP use. There were no significant differences in the populations captured by the simple versus alternative measures of motivation and access for VMMC, male condoms or PrEP, indicating that the simplest methods of defining motivation and access are sufficient to populate the HPC framework main bars.

Although Schaefer *et al*., and conclusions of the Harare workshop, argued that motivation and access are likely to be highly correlated^14^ and the order of the cascade framework will not make much difference to programmatic decision making^6^, swapping the order of the framework could mean that there are smaller drops offs observed in the cascade and therefore the areas identified as having the most potential to be improved by interventions could vary depending on the order of motivation and access. In this analysis, swapping the order of the motivation and access bars did not improve the information gained (i.e., larger gaps from which to identify barriers as targets for interventions) from the gaps between bars, and in the case of VMMC, actually meant less information about gaps in motivation could be gained from the cascade framework as most people were lost from the cascade in the first (access) bar.

Most barriers in the HPC were associated with a lack of effective use, supporting the theory behind the development of the framework in which the barriers were selected following preliminary analysis, literature review and behavioural theory^6,29^. These barriers identified through quantitative analyses are also supported by qualitative findings carried out within the same population^30,31^. Barriers to VMMC identified through qualitative focus groups and individual interviews included pain, fear of side effects and health consequences which pertain to the perceived consequences barrier of the cascade^31^. Barriers to PrEP use identified through these activities included a lack of knowledge and availability; barriers which were also indicated from the cascade analysis^32^.

### Weaknesses of the cascade framework

Non-trivial proportions of individuals reported both successful attainment of steps in the cascade and a barrier to that step, for example, being motivated to use a prevention method and also reporting a barrier to motivation to use that prevention method. For those who reported using a prevention method and still report barriers to use, it can be interpreted that these people had sufficient capacity to use the prevention method that these barriers did not prevent them from using the prevention method of interest. Additionally, for the motivation bar, if people reported wanting to use a prevention method then any reported barriers to motivation are not sufficient to prevent motivation. However, the definition of access used in the cascade – knowing somewhere to access a prevention method – does not directly preclude the barriers to access in the cascade, such as affordability. The current definition of the access bar means individuals could be classified as attaining access (main bar) who still have barriers to access which prevent them actually accessing a prevention method. Further work to assess variation in overlaps by strength of views on the main bars and sub-bars could also help to improve definitions of the main and sub-bars.

Effective prevention method use is challenging to measure, particularly with male condoms and PrEP. In absence of laboratory testing for the presence of PrEP drugs, which is impractical and expensive for large scale application of the prevention cascade, all measures of effective use of a prevention method rely on self-report which is subject to social desirability bias. The UNAIDS version of the HIV prevention cascade defines a focus population, similar to the LWG/Schaefer *et al*. priority population, and then reports reach/coverage uptake/use and correct/consistent use^16^. Taking into account consistent use is important when considering the ultimate goal of preventing acquisition of HIV but is difficult to measure using cross sectional survey data. Changing the definition of male condom use produced significantly different HIV prevention cascades for the same population.

Ideally, effective use should be consistent use; however, this also needs to be balanced with the practicality of collecting data on consistency of prevention method use especially when adding questions to existing surveys and acknowledge that HIV risk can change rapidly such as through life course events.

Most of the explanatory barriers are clearly defined, however some barriers encompass several variables which could contribute to that barrier. This particularly affects ‘lack of skills’ which could include social skills to negotiate prevention method use but also practical skills to actually use or adhere to prevention. Combining these aspects limits the insight which could be gained from the framework about potential targets for interventions. More granular information could be obtained if these were separated. There may be some populations in which male condoms have been widely available for a long time. In these cases, it may be appropriate to assume that male condoms are always available, and nobody experiences the lack of availability barrier, particularly when the space or time available to ask questions on explanatory barriers are limited.

This HPC framework was designed to be used for individual and combination prevention. When addressing combination prevention, it is possible to present an overall view of motivation, access, and use of multiple prevention methods in the priority population. However, including information about the explanatory barriers to all prevention methods of interest is complicated and nuanced. There are multiple options for how the bars could be populated, such as barriers reported across all prevention methods, or barriers to use of the prevention method furthest along the prevention cascade. However, this does not provide as complete a picture of explanatory barriers as focusing on individual prevention methods. Garnett *et al*. suggested an alternative approach to populating combination prevention cascades in which an HPC is constructed for the most widely used prevention method first, then for the next most widely used method for those not using the first method^5^. Although this approach is less succinct, it may reveal information on sub-bars for each prevention method.

### Limitations of this validation exercise

This validation exercise was limited by use of self-reported cross-sectional data, which may lead to underestimation of the size of the priority population and overestimation of levels of prevention method use. 23% of those eligible to complete the individual survey did not participate and 5% of participants did not consent to PITC or DBS collection which may introduce non-response bias to the population. It was not possible to address effective use of prevention method and the effect on HIV incidence, however multiple prevention methods have been proven to reduce HIV acquisition in other studies. When used correctly, condoms are highly effective in preventing transmission of HIV, giving an estimated reduction in transmission of 90-95%^33,34^. Voluntary medical male circumcision (VMMC) reduces HIV acquisition in men by between 53% and 60%^35–37^. The Zimbabwean national VMMC programme aims to reach 80% coverage of males aged 15-29 years by 2021 to reduce HIV incidence^38^. Clinical trials of oral pre-exposure prophylaxis (PrEP) have demonstrated its efficacy in preventing acquisition of HIV infection. Reported results vary, probably explained by differences in adherence^39–41^ - with good adherence the effectiveness of oral PrEP was as high as 90%^42^. No validation was carried out for applying the cascade framework to use of female condoms, although the prevention cascade for these shows that motivation and use of these is very low.

Validation of the HPC framework applied to PrEP use was limited due to very small numbers of individuals reporting awareness of or use of PrEP, which only very recently became available in Zimbabwe^43^. Motivation to use and access to PrEP were collinear with PrEP use. Although this demonstrates that those who reported PrEP use were motivated to use and did have access to PrEP, further analysis in populations with higher levels of PrEP use is required to confirm these associations. The association of explanatory barriers with PrEP use could not be tested due to such small numbers reporting the outcome.

Being motivated to have and reporting access to VMMC were associated with reduced odds of having VMMC, which is the opposite to what would be expected from the cascade framework. 95% of people reporting VMMC responded that they were not motivated to get VMMC or did not know somewhere to access VMMC if they did want to access it. If this issue was not considered when populating the cascade, for VMMC or other prevention methods, it could appear as if more people were using a prevention method than were actually motivated to use it or had access to it. Other studies which have applied the HIV prevention cascade have not reported or explored this, however, most have used different endpoint measures of the cascade. Hensen et al used the HPC framework to identify gaps to increase coverage of VMMC services in Zambia^44^, but the endpoint of the HPC framework was perceived service availability rather than actual uptake of VMMC. Some individuals reporting VMMC may have had VMMC when they were much younger, such as at school or as decided by their parents. Analysis of longitudinal data would be required to assess whether or not the men who have not yet taken up VMMC but do report motivation and access are more likely to do so compared to those who do not report motivation and access.

This validation exercise has only been tested in one population in Manicaland, east Zimbabwe. Further work using other populations would add to the credence of the HIV prevention cascade as a simple and effective way of understanding prevention method use. There were some explanatory barriers which were not associated with reduced odds of prevention method use, such as access related barriers to condom use, possibly because condoms are so widely available and accessible.

However, associations may be significant in other populations, and, therefore, the absence of associations within this population is not sufficient to warrant recommending removing these explanatory barriers from the cascade framework. The priority population chosen was a broad age range to maximise the sample size for analysis. Qualitative data suggest differences in attitudes to prevention method use in older versus younger people, highlighting the important of choosing relevant priority populations within appropriate age ranges^31^.

### Recommendations for the population of and structure of the cascade framework

Based on this analysis, we recommend the following updates to the LWG/Schaefer *et al*. HIV prevention cascade:

1. It should be assumed that all individuals reporting prevention method use are motivated and have access to that prevention method
2. Where possible, the definition of the access bar should not include anyone who reports any barriers to access
3. Motivation and access should remain as currently ordered, with the exception of populations where access is close to 100%. In this case, more information may be gained from putting the access bar first to maximise the gaps in the cascade and aid identification of targets for interventions, but any reported access related barriers should still be assessed.
4. The lack of skills explanatory barrier should be split into 2 separate barriers: a lack of social skills and a lack of practical skills
5. Where possible, quantitative analysis should be combined with qualitative analysis to understand barriers to use of prevention methods, especially in populations where awareness and use of a particular prevention method is very low.

### Applying the HIV Prevention Cascade

Overall, it is possible to collect data as part of routine population surveys to populate the HIV prevention cascade. However, this required inclusion of specific questionnaire modules and this type of data may not be available without the specific questionnaires through which this can be collected. Using Demographic and Health Survey data, it could be possible to populate the main bars of the cascade and thus generate a basic cascade used for high level monitoring and evaluation purposes.

This could mimic the success of the treatment cascade in allowing identification of the gaps in the cascade, monitoring these over time and making comparison across and within countries. Where large gaps are found, this could be complemented with surveys to specifically explain these gaps and thus find appropriate targets for interventions.

The UNAIDS version of the cascade is intended to be populated primarily using programme data that are already routinely available^16^. The main difference between this and the LWG/Schaefer cascade is that lack of motivation is not acknowledged within the UNAIDS cascade framework as an obstacle to use of prevention methods. Although gaps in programme coverage and use of prevention methods can be identified using the UNAIDS framework, when explaining these gaps and identifying relevant interventions, distinguishing motivation and demand-related factors from access-related explanatory factors is difficult. As demonstrated in this analysis, motivation is strongly associated with prevention method use and reporting barriers to motivation is associated with reduced odds of using a prevention method, supporting the importance of addressing this step of the cascade to increasing prevention method use even in a scenario of very high access to primary prevention. Other efforts to populate this cascade framework, such as that looking at condom use in young women who sell sex in Zimbabwe, did not have data available on motivation to use condoms and so had to use a proxy measure of knowledge about condom efficacy^21^.

For a standardised approach for comparing countries, there is a strong preference and benefit to using one version of the cascade framework in order to mimic the success of the treatment cascade and allow comparison across populations. As Auerbach *et al*. propose, a solution could be to use a 2-step process involving two cascades^17^. The first step would use the UNAIDS framework, leveraging commonly available programmatic data to populate the cascade and identify gaps. Once the gaps are identified, the LWG/Schaefer *et al*. cascade can be used to further understand the demand side of prevention method use and reasons for gaps in the cascade using the hypothesised explanatory sub-bars. Given the demonstrated importance of motivation within the cascade framework, adding a question on motivation wherever possible, such as to routine surveys, would provide valuable information on the gaps in individual level motivation to use primary prevention methods. The latest WHO Strategic Information guidelines on HIV prevention emphasise demand-led referrals to primary prevention services^45^. Monitoring of this could mean that data on whether or not individuals seen in routine programmes want (are motivated) to use a prevention method would need to be collected in programme records, thus making is possible to measure the LWG/Schaefer HPC using programme data. However, these records would still only capture data on people who access or are reached by the programme.

Schaefer *et al*. note that the LWG/Schaefer proposed version of the cascade could be applied to mathematical modelling to predict the infections averted by current HIV prevention use and predict the impact on HIV incidence of reducing barriers within the cascade^6^, as also suggested by Auerbach *et al*.^17^, and demonstrated by Pickles *et al* using data from this study to parameterise the model using this version of the cascade framework^46^.

### Final recommended formulation of the HIV Prevention Cascade

The final proposed formulation of the HIV prevention cascade is illustrated for men’s use of male condoms in Figure 10. Depending on the availability of data, the cascade can be populated as the full cascade including explanatory barriers to each step, or just as the main bars of the cascade whereby the difference in the bars indicates the gaps where people are being lost from the cascade. In the latter case, once these gaps are identified, further work can be done to establish the cause of these gaps.

### Steps to populating the HIV Prevention Cascade

The following steps should be taken to populate this HIV prevention cascade:

1) **Priority population:** Establish the priority population as a population that would benefit from using HIV prevention according to the population or research question of interest
2) **Main bars:**

**a. Use:** Calculate the number within the priority population using the HIV prevention method of interest according to the chosen definition of effective use. Suggested definitions are listed in Table 7.
**b. Motivation:** Calculate the number within the priority population who are motivated to use the HIV prevention method of interest according to the chosen definition of motivation. Suggested definitions are listed in Table 7. Recode all individuals who are reporting use but not motivation to be motivated.
**c. Access:** Calculate the number within the priority population who are motivated to use and report access to the HIV prevention method of interest according to the chosen definition of access. Suggested definitions are listed in Table 7. Recode all individuals who are reporting use but not access to have access. Where data is available, recode all individuals reporting motivation and at least one barrier to access as not having access.
**d. Calculate motivation, access, and use as proportions:** Each main bar of the cascade is presented as a proportion with the calculations:

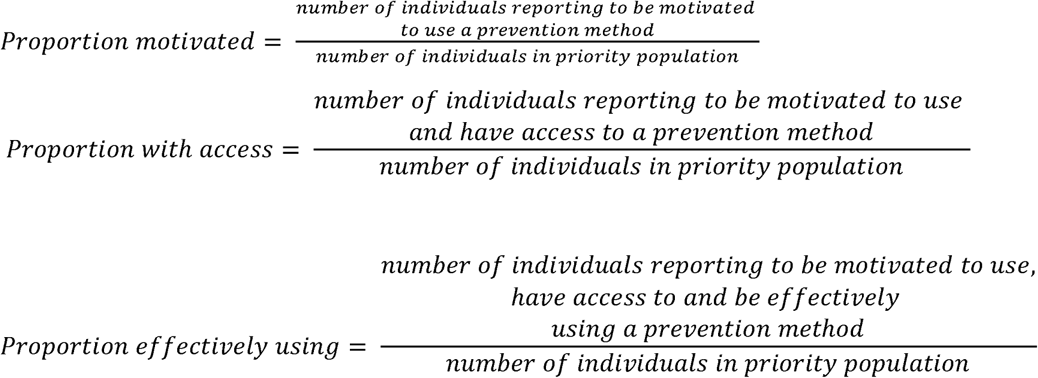

95% confidence intervals for each main bar proportion can be calculated and displayed around the main bars of the HPC framework.

3) **Explanatory sub bars:** Where data are available explanatory sub bars for each step can be populated using suggested definitions in Table 8. The explanatory sub bars should be limited to those falling within the gaps between each of the main bars in the cascade.

Motivation related sub-bars should only be experienced by those who are in the priority population but unmotivated. Access related sub-bars should only be experienced by those in the priority population who are motivated but do not report access. Effective use sub-bars should only be experienced by those in the priority population who are motivated and have access but do not report using the prevention method of interest.

4) **Combination prevention:** Where data are available, the measures of individual prevention method motivation, access and use can be combined to produce combination prevention cascades. Depending on the population of interest and research questions, criteria for combination motivation, access or use can either be:

**a. Using at least one prevention method –** bars for combination use should be created where individual meeting the criteria for the respective bar for at least one prevention method fall within that bar
**b. Using multiple prevention methods at the same time -** bars for combination use should be created where individuals meeting the criteria for the respective bar for at all prevention methods of interest fall within that bar

Main bars only should be populated for combination prevention cascades. If gaps are identified from this analysis, the cascades should be split into individual prevention methods and at this point explanatory sub bars should be populated to understand the barriers relevant to each prevention method. At this point, common barriers across prevention methods within the priority population could be identified.

1. **Comparison to national/international targets –** calculate the percentage of the priority population who report motivation, the percentage of those motivated who report access and the percentage of those motivated and with access who report effect use. Compare these with the 90-90-90 equivalent targets where available, such as those set out in the UNAIDS HIV Prevention 2025 Road Map^47,48^.

## Conclusions

Overall, it was possible to measure the HPC for individual and combination prevention using the questionnaires module designed for the Manicaland pilot survey. It has now been established, through our work, that it is possible to populate an HPC framework with data collected in a general population survey using questionnaire modules developed to facilitate this process. Through a combination of literature review, social and behavioural theory, and evaluation using real world data, a general consensus has been reached on what is important to include in a basic HIV prevention cascade and how such a framework can be used to support HIV prevention efforts. If sufficiently evaluated and promoted, the HIV prevention cascade can contribute to improvements in HIV prevention service provision and help to bring about necessary reductions in HIV incidence to end the HIV/AIDS epidemic as a global public health threat.

## Data Availability

Data access statement
Due to the sensitive nature of data collected, including information on HIV status, treatment and sexual risk behaviour, the Manicaland Centre for Public Health does not make full analysis datasets publicly available. Summary datasets of household and background sociodemographic individual questionnaire data, covering rounds 1-8 (1998-2021), are publicly available for download via the Manicaland Centre for Public Health website here - http://www.manicalandhivproject.org/data-access.html. Quantitative data used for analyses produced by the Manicaland Centre for Public Health are available on request following completion of a data access request form here - http://www.manicalandhivproject.org/data-access.html. Additionally, summary HIV incidence and mortality data spanning rounds 1-6 (1998-2013), created in collaboration with the ALPHA Network are available via the DataFirst Repository here - https://www.datafirst.uct.ac.za/dataportal/index.php/catalog/ALPHA/about

## Acknowledgements

We are very grateful to the research team working at the Manicaland Centre for Public Health, as well as to the study participants in Manicaland, for their contribution to this study.

SG, TBH, CN, JI-E and LM conceptualised the study. CN, BT, RM, and PM-M had major roles in collection and management of data used in this study. LM analysed the data. LM, SG and JI-E contributed to interpretation of results. LM wrote the initial report which was reviewed and revised by all co-authors. All authors had final responsibility for the decision to submit for publication.

## Data access statement

Due to the sensitive nature of data collected, including information on HIV status, treatment and sexual risk behaviour, the Manicaland Centre for Public Health does not make full analysis datasets publicly available. Summary datasets of household and background sociodemographic individual questionnaire data, covering rounds 1-8 (1998-2021), are publicly available for download via the Manicaland Centre for Public Health website here - http://www.manicalandhivproject.org/data-access.html. Quantitative data used for analyses produced by the Manicaland Centre for Public Health are available on request following completion of a data access request form here - http://www.manicalandhivproject.org/data-access.html. Additionally, summary HIV incidence and mortality data spanning rounds 1-6 (1998-2013), created in collaboration with the ALPHA Network are available via the DataFirst Repository here - https://www.datafirst.uct.ac.za/dataportal/index.php/catalog/ALPHA/about

---

Table S1 - Male condom cascade main bar and sub bar definitions Table S2 - PrEP cascade main bar and sub bar definitions

Table S3 - VMMC cascade main bar and sub bar definitions

Table S4 - Proposed revised definitions of each main bar of the HIV prevention cascade

Table S5 - Proposed revised definitions of each explanatory sub-bar in the HIV prevention cascade

