## Supplementary material for "Appraising the HIV Prevention Cascade methodology to improve HIV prevention targets: Lessons learned from a general population pilot study in east Zimbabwe": Table S1

Table S1 - Male condom cascade main bar and sub bar definitions

| Prevention Cascade Bar/Sub-bar | Criteria to be met for an individual to experience a bar/sub-bar |
| --- | --- |
| Motivation: |  |
| Wants to use prevention method | **Simple measure:**  Regular partners: Wants to use male condoms with regular partner if they were freely accessible  Non-regular partners:  Wants to use male condoms with non-regular partner if they were freely accessible  Regular and non-regular: if respondent has both regular and non-regular they need to be motivated for both of these partners, so combine the two measures above.  **Alternative measure:**  Above plus  Regular partners:   - respondent defines this want as ‘definitely’ or ‘probably’ from a scale of ‘definitely’, ‘probably’, ‘no opinion’, ‘probably not’ ‘definitely not’   Non-regular partners:   - respondent defines this want as ‘definitely’ or ‘probably’ from a scale of ‘definitely’, ‘probably’, ‘no opinion’, ‘probably not’ ‘definitely not’ |
| Barriers to motivation: |  |
| Knowledge | At least one of:   - Respondent has not heard of male condoms, either spontaneously or when probed. - Respondent responds that male condoms reduce a person’s risk of getting HIV infection by <80%^25^ |
| Risk perception | Respondent perceives no or small future risk of HIV infection in the next 12 months |
| Perceived negative consequences | At least one of:   - Respondent is discouraged from using male condoms due to own sexual pleasure |
| Social acceptability | At least one of:   - Respondent is discouraged from using male condoms due to making them feel responsible or ashamed - Respondent reported views of any of religious leaders discouraged male condom use - Respondent reported views of any of parents/family elders discouraged male condom use - Respondent reported friends/community thinking they have HIV discouraged male condom use - Respondent reported friends/community views discouraged male condom use - Respondent disagrees it is acceptable for a husband and wife to use condoms (i) always, or (ii) if at least one of them is HIV+, or (iii) if one spouse has other partners, or (iv) if one spouse has an STI. |
| Access: |  |
| Can access prevention method | **Simple measure:**  Respondent responds if/when they want to use male condoms they know a place where they can get them  (*Plus, those who are effectively using but do not meet access criteria*)  **Alternative measure:**   - Respondent responds if/when they want to use male condoms they know a place where they can get them and this access is ‘very easy’ or ‘easy’ or ‘neither easy nor difficult’ on a scale of ‘very easy’, ‘easy’, ‘neither easy nor difficult’, ‘difficult’ or ‘very difficult’. |
| Barriers to access: |  |
| Availability | Respondent doesn’t know a place where male condoms are available. |
| Acceptable provision | At least one of:   - Respondent responds that it is impractical or unsuitable to access male condoms due to lack of privacy/confidentiality - Respondent responds that it is impractical or unsuitable to access male condoms because embarrassed to go/ask - Respondent did not use male condoms because of judgemental staff/stigma |
| Easy access | At least one of:   - Respondent reports limited opening hours make it impractical to access male condoms - Respondent rates access as ‘difficult’ or ‘very difficult’ to access male condoms if/when you they want to use them (unless using this question as the Simple measure for the main access bar) on a scale of ‘very easy’, ‘easy’, ‘neither easy nor difficult’, ‘difficult’ or ‘very difficult’. - Respondent feels unable to access male condoms from any of:   - Health clinic   - Community based distributor   - Bars or beer halls   - Shops   - Sexual partner   - Friends |
| Affordability | - Reports high costs making it impractical to access male condoms |
| Effective use: |  |
| Effectively using prevention method  *NB – effective use definition is dependent on period of interest of risk and priority population* | **Simple measure:**   - Respondent reports using male condoms as HIV prevention method now     *If lifetime partners or other time period (depending on priority population and period of risk focusing on)*  Regular partner   - Used male condoms for all sexually active/>1 year every time they have had sex with a regular partner   Non-regular partner   - Used male condoms for all sexually active/>1 year they have had sex with a non-regular partner   Or   - Reports using male condoms throughout last sex |
| Barriers to effective use: |  |
| Skills | At least one of:  Regular partners:   - Respondent is not able to refuse sex with partner if regular partner does not want to use male condoms - Respondent is not able to discuss using male condoms with regular partner   Non-regular partners:   - Respondent is not able to refuse sex with partner if non-regular partner does not want to use male condoms - Respondent is not able to discuss using male condoms with non-regular partner   All *(unless these are being used in definition of effective use)*   - Respondent has not received instructions or counselling on how to use male condoms - Respondent does not replace male condom if it breaks |
| Self-efficacy | At least one of:   - Respondent not confident can use male condoms if wanted to - Respondent not confident can use male condoms even if they have to use them every time - Respondent not confident can use male condoms even if partner dislikes/disapproves - Respondent not confident can use male condoms even if my friends disapprove - Respondent not confident can use male condoms even if parents and family elders disapprove |
| Partner | At least one of:  Regular partner   - Regular partner does/would disapprove of using male condoms   Non-regular partner   - Non-regular partner does/would disapprove of using male condoms   All   - Respondent was discouraged from using male condoms because partner(s) will think respondent has HIV - Respondent was discouraged from using male condoms because partner will think respondent has other partners - Respondent was discouraged from using male condoms because of partner’s views - Respondent is discouraged from using male condoms due to partner’s sexual pleasure |
