## Supplementary material for "Appraising the HIV Prevention Cascade methodology to improve HIV prevention targets: Lessons learned from a general population pilot study in east Zimbabwe": Table S2

Table S2 - PrEP cascade main bar and sub bar definitions

| Prevention Cascade Bar/Sub-bar | Criteria to be met for individual experience a bar/sub-bar |
| --- | --- |
| Motivation: |  |
| Wants to use prevention method | **Simple measure:**  At least one of:   - Respondent plans to start using PrEP - Respondent wants to use PrEP if it was freely accessible   **Alternative measure:**   - Respondent definitely or probably wants to use PrEP if it was freely accessible from a scale of ‘definitely’, ‘probably’, ‘no opinion’, ‘probably not’ ‘definitely not’ |
| Barriers to motivation: |  |
| Knowledge | At least one of:   - Respondent has not heard about PrEP - Respondent does not think PrEP works - Respondent thinks PrEP reduces risk of HIV infection by <80% |
| Risk perception | Respondent perceives no or small future risk of HIV infection in the next 12 months |
| Perceived consequences | At least one of:   - Respondent stopped PrEP/did not use PrEP due to experiencing side effects - Respondent discouraged from using PrEP due to possible side effects |
| Social acceptability | At least one of:   - Respondent is discouraged from using PrEP due to making them feel responsible or ashamed - Respondent reported views of any of parents/family elders discouraged PrEP use - Respondent reported views of any of religious leaders discouraged PrEP use - Respondent reports friends (or their partners) are not using PrEP |
| Access: |  |
| Can access prevention method | **Simple measure:**  At least one of:   - Respondent knows a place to access PrEP   **Alternative measure:**   - Respondent reports it is easy or very easy to access PrEP services if/when they want to use it   (*Plus, those who are effectively using but do not meet access criteria*) |
| Barriers to access: |  |
| Availability | At least one of:   - Respondent stopped using PrEP/does not use PrEP due to stock out drugs at clinics - Respondent has never been offered PrEP (depending on priority population) |
| Acceptable provision | At least one of:   - Respondent stopped PrEP/did not use PrEP due to lack of confidentiality - Respondent stopped PrEP/did not use PrEP due to judgemental staff/stigma - Respondent discouraged from accessing PrEP services due to a lack of privacy/confidentiality - Respondent responds that it is impractical or unsuitable to access PrEP services because embarrassed to go/ask |
| Easy access | At least one of:   - Respondent reports not using PrEP due to limited opening hours - Respondent reports not using PrEP due to distance/travel difficulties - Respondent reports it is impractical or unsuitable to access PrEP services because of limited opening hours - Respondent reports it is difficult or very difficult to access PrEP services if/when they want to use it (depending on if this question is used in main bar definition of having access) on a scale of ‘very easy’, ‘easy’, ‘neither easy nor difficult’, ‘difficult’ or ‘very difficult’. |
| Affordability | At least one of:   - Respondent stopped using/did not use PrEP due to high costs - Respondent discouraged from using PrEP due to high costs involved - Respondent reports it is impractical or unsuitable to access PrEP services because of high costs |
| Effective use: |  |
| Effectively using prevention method | **Simple measure:**  At least one of:   - Respondent self-reports taking PrEP currently - Respondent self-reports using PrEP as an HIV prevention method currently   **Alternative measures:**   - Laboratory testing for PrEP adherence (if available) - Respondent reports having taken PrEP every day or most days in the last month |
| Barriers to effective use: |  |
| Skills | At least one of:   - Respondent did not use PrEP/stopped using PrEP due to forgetting to take pills - Respondent did not use PrEP/stopped using PrEP due to running out of pills - Respondent has not received instruction/counselling on how to use PrEP - Respondent does not always or mostly does not take PrEP with a meal - Respondent is not able to discuss taking PrEP with partner |
| Self-efficacy | At least one of:  (With disagreement on a scale of ‘strongly disagrees’ ‘disagrees’, ‘neutral’, ‘agree’, ‘strongly agree’)   - Respondent (strongly) disagrees that they are confident to use PrEP if they wanted to - Respondent (strongly) disagrees that they are confident to use PrEP even if they have to take it every day - Respondent (strongly) disagrees that they are confident to use PrEP even if always taken after a meal - Respondent is discouraged from using PrEP even if friends disapprove - Respondent is discouraged from using PrEP even if family elders/parents disapprove - Respondent (strongly) disagrees that they are confident to use PrEP even if community would think they have HIV - Respondent is discouraged from using PrEP because it is inconvenient to take pills daily |
| Partner | At least one of:  (With disagreement on a scale of ‘strongly disagrees’ ‘disagrees’, ‘neutral’, ‘agree’, ‘strongly agree’)   - Respondent (strongly) disagrees that they are confident in using PrEP even if they have to hide it from partner - Respondent reports partner would disapprove if they used PrEP - Respondent does not use/stopped using PrEP due to sexual partner disapproval |
