## Supplementary material for "Appraising the HIV Prevention Cascade methodology to improve HIV prevention targets: Lessons learned from a general population pilot study in east Zimbabwe": Table S3

Table S3 - VMMC cascade main bar and sub bar definitions

| Prevention Cascade Bar/Sub-bar | Criteria to be met for individual experience a bar/sub-bar |
| --- | --- |
| Motivation: |  |
| Wants to use prevention method | **Simple measure:**  At least one of:   - Respondent reports they would get VMMC if it was freely accessible - Respondent plans to get VMMC   **Alternative measure:**   - Respondent reports they would definitely or probably get VMMC if it was freely accessible from a scale of ‘definitely’, ‘probably’, ‘no opinion’, ‘probably not’ ‘definitely not’ |
| Barriers to motivation: |  |
| Knowledge | At least one of:   - Respondent has not heard of VMMC - Respondent has not been offered VMMC - Respondent thinks reduction of risk in infection offered by VMMC is less than 50%^26^ |
| Risk perception | Respondent perceives no or small future risk of HIV infection in the next 12 months |
| Perceived consequences | At least one of:   - Respondent is discouraged from having VMMC due to pain - Respondent is discouraged from having VMMC because operation cannot be reversed - Respondent is discouraged from having VMMC because of own sexual pleasure |
| Social acceptability | At least one of:   - Respondent is discouraged from having VMMC because of views of religious leader - Respondent is discouraged from having VMMC because of views of parents or family elder - Respondent reports friends (or their partners) have not had VMMC |
| Access: |  |
| Can access prevention method | **Simple measure:**  At least one of:   - Respondent knows a place where they could get VMMC if they wanted to get it   (*Plus, those who are effectively using but do not meet access criteria*)  **Alternative measure**  Respondent knows a place where it is not difficult or very difficult to get VMMC on a scale of ‘very easy’, ‘easy’, ‘neither easy nor difficult’, ‘difficult’ or ‘very difficult’. |
| Barriers to access: |  |
| Availability | Respondent reports that it is difficult or very difficult to access VMMC services if they wished to on a scale of ‘very easy’, ‘easy’, ‘neither easy nor difficult’, ‘difficult’ or ‘very difficult’. |
| Acceptable provision | At least one of:   - Respondent responds that it is impractical or unsuitable to access VMMC services because of lack of privacy/confidentiality - Respondent responds that it is impractical or unsuitable to access VMMC services because healthcare workers are female |
| Easy access | At least one of:   - Respondent reports it is impractical or unsuitable to access VMMC services due to distance/travel difficulties - Respondent reports it is impractical or unsuitable to access VMMC services due to limited opening hours |
| Affordability | At least one of:   - Respondent reports it being impractical or unsuitable to access VMMC services due to high cost including loss of income - Respondent reports it being impractical or unsuitable to access VMMC services due to inability to work during/after procedure |
| Effective use: |  |
| Effectively using prevention method | **Simple measure:**  At least one of:   - Respondent has had full medical circumcision - Respondent has had full medical and traditional/religious circumcision   **Alternative measure:**  Clinic confirmed VMMC record if available |
| Barriers to effective use: |  |
| Skills | Respondent cannot discuss getting VMMC with partner |
| Self-efficacy | At least one of:  (With disagreement on a scale of ‘strongly disagrees’ ‘disagrees’, ‘neutral’, ‘agree’, ‘strongly agree’)   - Respondent (strongly) disagrees that they are confident they can get VMMC if they want to - Respondent (strongly) disagrees that they are confident they can get VMMC even if friends disapprove - Respondent (strongly) disagrees that they are confident they can get VMMC even if parents and family elders disapprove |
| Partner | At least one of:  (With disagreement on a scale of ‘strongly disagrees’ ‘disagrees’, ‘neutral’, ‘agree’, ‘strongly agree’)   - Respondent reports that partner would disapprove if they had VMMC - Respondent is discouraged from having VMMC because of partner’s sexual pleasure - Respondent (strongly) disagrees that they are confident they can get VMMC even if partner disapproves - Respondent is discouraged from having VMMC because of partner’s views |
