## Supplementary material for "Appraising the HIV Prevention Cascade methodology to improve HIV prevention targets: Lessons learned from a general population pilot study in east Zimbabwe": Table S4

Table S4 - Proposed revised definitions of each main bar of the HIV prevention cascade

| **Prevention method** |  | **Prevention Cascade Domain** |  | **Measure to populate bar** |
| --- | --- | --- | --- | --- |
| **Male condoms** |  | Motivation |  | Regular partners: Wants to use male condoms with regular partner if they were freely accessible  Non-regular partners: Wants to use male condoms with non-regular partner if they were freely accessible Regular and non-regular: if respondent has both regular and non-regular they need to be motivated for both of these partners, so combine the two measures above. |
|  |  | Access |  | Respondent responds if/when they want to use male condoms they know a place where they can get them  (plus those who are effectively using but do not meet access criteria) **AND**  Not reporting any barriers to access to male condoms |
|  |  | Use |  | Respondent reports using male condoms at last sex as HIV prevention method now (if available, with the relevant type of partner (regular/non-regular) |
| **PrEP** |  |  |  |  |
|  |  | Motivation |  | At least one of: - Respondent wants to use PrEP if it was freely accessible  **IF CAPACITY FOR SECOND QUESTION**  - Respondent plans to start using PrEP |
|  |  | Access |  | Respondent responds if/when they want to use PrEP they know a place where they can get them  (plus those who are effectively using but do not meet access criteria) **AND**  Not reporting any barriers to access to PrEP |
|  |  | Use |  | Respondent reports full medical male circumcision  **AND IF AVAILABLE FOR PRIORITY POPULATION** Clinic confirmed PrEP use or adherence testing |
| **VMMC** |  | Motivation |  | At least one of:  - Respondent reports they would get VMMC if it was freely accessible  **IF CAPACITY FOR SECOND QUESTION** - Respondent plans to get VMMC |
|  |  | Access |  | Respondent responds if/when they want to use VMMC they know a place where they can get them  (plus those who are effectively using but do not meet access criteria) **AND**  Not reporting any barriers to access to VMMC |
|  |  | Use |  | Respondent reports full medical male circumcision  **AND IF AVAILABLE FOR PRIORITY POPULATION** Clinic confirmed VMMC, or respondent shows VMMC certificate |
