## Supplementary material for "Appraising the HIV Prevention Cascade methodology to improve HIV prevention targets: Lessons learned from a general population pilot study in east Zimbabwe": Table S5

Table S5 - proposed revised definitions of each explanatory sub-bar in the HIV prevention cascade

| **Prevention method** |  | **Prevention Cascade Domain** |  |  |  |
| --- | --- | --- | --- | --- | --- |
|  |  |  |  | **Barrier** | **Definition** |
| **Male condoms** |  | Motivation |  | Lacks knowledge | At least one of:  - Respondent has not heard of male condoms, either spontaneously or when probed.  - Respondent responds that male condoms reduce a person’s risk of getting HIV infection by <80% |
|  |  |  |  | Lacks risk perception | Respondent perceives no or small future risk of HIV infection in the next 12 months |
|  |  |  |  | Perceives consequences | At least one of:  - Respondent is discouraged from using male condoms due to loss of own sexual pleasure |
|  |  |  |  | Social unacceptability | At least one of:  - Respondent is discouraged from using male condoms due to making them feel irresponsible or ashamed  - Respondent reported views of any of religious leaders discouraged male condom use  - Respondent reported views of any of parents/family elders discouraged male condom use - Respondent reported friends/community thinking they have HIV discouraged male condom use - Respondent reported friends/community views discouraged male condom use - Respondent disagrees it is acceptable for a husband and wife to use condoms (i) always, or (ii) if at least one of them is HIV+, or (iii) if one spouse has other partners, or (iv) if one spouse has an STI. |
|  |  | Access |  | Lack of availability | Respondent doesn’t know a place where male condoms are available.  *Depending on context, there may be populations where male condoms are widely available and so it could be assumed that male condoms are always available, and nobody experiences this barrier.* |
|  |  |  |  | Lack of affordability | Respondent reports high costs making it impractical to access male condoms |
|  |  |  |  | Lack of easy access | At least one of:  - Respondent reports limited opening hours make it impractical to access male condoms  - Respondent rates access as ‘difficult’ or ‘very difficult’ to access male condoms if/when you they want to use them (unless using this question as the Simple measure for the main access bar) - Respondent feels unable to access male condoms from any of:  - Health clinic  - Community based distributor  - Bars or beer halls  - Shops   - Sexual partner  - Friends |
|  |  |  |  | Lack of acceptable provision | At least one of:  - Respondent responds that it is impractical or unsuitable to access male condoms due to lack of privacy/confidentiality  - Respondent responds that it is impractical or unsuitable to access male condoms because embarrassed to go/ask - Respondent did not access male condoms because of judgemental staff/stigma |
|  |  | Use |  | Lack of self-efficacy | At least one of:  - Respondent not confident can use male condoms if wanted to  - Respondent not confident can use male condoms if they have to use them every time - Respondent not confident can use male condoms if partner dislikes/disapproves - Respondent not confident can use male condoms if my friends disapprove - Respondent not confident can use male condoms if parents and family elders disapprove |
|  |  |  |  | Lack of social skills | At least one of:  Regular partners:  - Respondent is not able to refuse sex with partner if regular partner does not want to use male condoms  - Respondent is not able to discuss using male condoms with regular partner Non-regular partners:  - Respondent is not able to refuse sex with partner if non-regular partner does not want to use male condoms  - Respondent is not able to discuss using male condoms with non-regular partner |
|  |  |  |  | Lack of practical skills | At least one of:  - Respondent has not received instructions or counselling on how to use male condoms  - Respondent does not replace male condom if it breaks |
|  |  |  |  | Partner | At least one of: Regular partner - Regular partner does/would disapprove of using male condoms Non-regular partner - Non-regular partner does/would disapprove of using male condoms  All - Respondent was discouraged from using male condoms because partner(s) will think respondent has HIV  - Respondent was discouraged from using male condoms because partner will think respondent has other partners - Respondent was discouraged from using male condoms because of partner’s views  - Respondent is discouraged from using male condoms due to partner’s sexual pleasure |
| **PrEP** |  | Motivation |  | Lacks knowledge | At least one of:  - Respondent has not heard about PrEP  - Respondent does not think PrEP works - Respondent thinks PrEP reduces risk of HIV infection by <80% |
|  |  |  |  | Lacks risk perception | Respondent perceives no or small future risk of HIV infection in the next 12 months |
|  |  |  |  | Perceives consequences | At least one of:  - Respondent stopped PrEP/did not use PrEP due to experiencing side effects - Respondent discouraged from using PrEP due to possible side effects |
|  |  |  |  | Social unacceptability | At least one of:  - Respondent is discouraged from using PrEP due to making them feel irresponsible or ashamed  - Respondent reported views of any of parents/family elders discouraged PrEP use  - Respondent reported views of any of religious leaders discouraged PrEP use  - Respondent reports friends (or their partners) are not using PrEP |
|  |  | Access |  | Lack of availability | At least one of: - Respondent stopped using PrEP/does not use PrEP due to stock out drugs at clinics - Respondent has never been offered PrEP |
|  |  |  |  | Lack of affordability | At least one of:  - Respondent stopped using/did not use PrEP due to high costs  - Respondent discouraged from using PrEP due to high costs involved  - Respondent reports it is impractical or unsuitable to access PrEP services because of high costs |
|  |  |  |  | Lack of easy access | At least one of:  - Respondent reports not using PrEP, or it is impractical or unsuitable to access PrEP due to limited opening hours  - Respondent reports not using PrEP due to distance/travel difficulties  - Respondent reports it is difficult or very difficult to accessing PrEP services if/when they want to use it (depending on if this question is used in main bar definition of having access) |
|  |  |  |  | Lack of acceptable provision | At least one of:  - Respondent stopped PrEP/did not use PrEP due to lack of confidentiality - Respondent stopped PrEP/did not use PrEP due to judgemental staff/stigma - Respondent discouraged from accessing PrEP services due to a lack of privacy/confidentiality - Respondent responds that it is impractical or unsuitable to access PrEP services because embarrassed to go/ask |
|  |  | Use |  | Lack of self-efficacy | At least one of:  (With disagreement on a scale of ‘strongly disagrees’ ‘disagrees’, ‘neutral’, ‘agree’, ‘strongly agree’) - Respondent (strongly) disagrees that they are confident to use PrEP if they wanted to  - Respondent is discouraged from using PrEP if friends disapprove - Respondent is discouraged from using PrEP if family elders/parents disapprove  - Respondent (strongly) disagrees that they are confident to use PrEP if community would think they have HIV |
|  |  |  |  | Lack of social skills | Respondent is not able to discuss taking PrEP with partner |
|  |  |  |  | Lack of practical skills | At least one of:  -Respondent is discouraged from using PrEP because it is inconvenient to take pills daily - Respondent (strongly) disagrees that they are confident to use PrEP if they have to take it every day (with disagreement on a scale of ‘strongly disagrees’ ‘disagrees’, ‘neutral’, ‘agree’, ‘strongly agree’) - Respondent did not use PrEP/stopped using PrEP due to forgetting to take pills  - Respondent did not use PrEP/stopped using PrEP due to running out of pills  - Respondent has not received instruction/counselling on how to use PrEP - Respondent does not always or mostly does not take PrEP with a meal |
|  |  |  |  | Partner | At least one of:  (With disagreement on a scale of ‘strongly disagrees’ ‘disagrees’, ‘neutral’, ‘agree’, ‘strongly agree’) - Respondent (strongly) disagrees that they are confident in using PrEP even if they have to hide it from partner - Respondent reports partner would disapprove if they used PrEP  - Respondent does not use/stopped using PrEP due to sexual partner disapproval |
| **VMMC** |  | Motivation |  | Lacks knowledge | At least one of:  - Respondent has not heard of VMMC  - Respondent has not been offered VMMC  - Respondent thinks reduction of risk in infection offered by VMMC is less than 50% |
|  |  |  |  | Lacks risk perception | Respondent perceives no or small future risk of HIV infection in the next 12 months |
|  |  |  |  | Perceives consequences | At least one of:  - Respondent is discouraged from having VMMC due to pain - Respondent is discouraged from having VMMC because operation cannot be reversed - Respondent is discouraged from having VMMC because of loss of own sexual pleasure |
|  |  |  |  | Social unacceptability | At least one of:  - Respondent is discouraged from having VMMC because of views of religious leader - Respondent is discouraged from having VMMC because of views of parents or family elder  - Respondent reports friends (or their partners) have not had VMMC |
|  |  | Access |  | Lack of availability | Respondent reports that it is difficult or very difficult to access VMMC services if they wished to |
|  |  |  |  | Lack of affordability | At least one of:  - Respondent reports it being impractical or unsuitable to access VMMC services due to high cost including loss of income  - Respondent reports it being impractical or unsuitable to access VMMC services due to inability to work during/after procedure |
|  |  |  |  | Lack of easy access | At least one of:  - Respondent reports it is impractical or unsuitable to access VMMC services due to distance/travel difficulties - Respondent reports it is impractical or unsuitable to access VMMC services due to limited opening hours |
|  |  |  |  | Lack of acceptable provision | At least one of:  - Respondent responds that it is impractical or unsuitable to access VMMC services because of lack of privacy/confidentiality  - Respondent responds that it is impractical or unsuitable to access VMMC services because healthcare workers are female |
|  |  | Use |  | Lack of self-efficacy | At least one of:  (With disagreement on a scale of ‘strongly disagrees’ ‘disagrees’, ‘neutral’, ‘agree’, ‘strongly agree’) - Respondent (strongly) disagrees that they are confident they can get VMMC if they want to - Respondent (strongly) disagrees that they are confident they can get VMMC if friends disapprove - Respondent (strongly) disagrees that they are confident they can get VMMC if parents and family elders disapprove |
|  |  |  |  | Lack of social skills | Respondent cannot discuss getting VMMC with partner |
|  |  |  |  | Partner | At least one of: - Respondent reports that partner would disapprove if they had VMMC - Respondent is discouraged from having VMMC because of partner’s sexual pleasure - Respondent (strongly) disagrees that they are confident they can get VMMC if partner disapproves (with disagreement on a scale of ‘strongly disagrees’ ‘disagrees’, ‘neutral’, ‘agree’, ‘strongly agree’) - Respondent is discouraged from having VMMC because of partner’s views |
